# The Health Effects of Climate Change: An Overview of Systematic Reviews

**DOI:** 10.1101/2020.09.29.20204123

**Authors:** Rhéa Rocque, Caroline Beaudoin, Ruth Ndjaboue, Laura Cameron, Louann Poirier Bergeron, Rose-Alice Poulin-Rheault, Catherine Fallon, Andrea C. Tricco, Holly O. Witteman

## Abstract

**Background:** Although many studies have explored the health impacts of climate change, a broader overview of research is needed to guide future research and action to mitigate and adapt to the health impacts of climate change.

**Methods:** We conducted an overview of systematic reviews of health impacts of climate change. We systematically searched the literature using a predefined search strategy, inclusion, and exclusion criteria. We included systematic reviews that explored at least one health impact of climate change. We organized systematic reviews according to their key characteristics, including geographical regions, year of publication and authors’ affiliations. We mapped the climate effects and health outcomes being studied and synthesized major findings.

**Findings:** We included ninety-four systematic reviews. Most were published after 2015 and approximately one fifth contained meta-analyses. Reviews synthesized evidence about five categories of climate impacts; the two most common were meteorological and extreme weather events. Reviews covered ten health outcome categories; the three most common were 1) infectious diseases, 2) mortality, and 3) respiratory, cardiovascular, cardiopulmonary or neurological outcomes. Most reviews suggested a deleterious impact of climate change on multiple adverse health outcomes, although the majority also called for more research.

**Interpretation:** Overall, most systematic reviews suggest that climate change is associated with worse human health. Future research could explore the potential explanations between these associations to propose adaptation and mitigation strategies and could include psychological and broader social health impacts of climate change.

**Funding:** Canadian Institutes of Health Research FDN-148426

## Introduction

The environmental consequences of climate change such as rising temperatures, more extreme weather events, and increased droughts and flooding are impacting human health and lives.^1,2^ Previous studies and reviews have documented the health impacts of climate change; however, they have focused on specific climate effects,^3,4^ health impacts,^5,6^ countries,^7–9^ or are no longer up to date.^10,11^ To guide future research and action to mitigate and adapt to the health impacts of climate change and its environmental consequences, we need a complete and thorough overview of the research already conducted. In this study, we aimed to develop such a synthesis of systematic reviews of health impacts of climate change. Our research objectives were to synthesize studies’ characteristics such as geographical regions, years of publication, and authors’ affiliations, to map the climate impacts, health outcomes, and combinations of these that have been studied, and to synthesize key findings.

## Methods

We applied the Cochrane method for overviews of reviews.^12^ This method is designed to systematically map the themes of studies on a topic and synthesize findings to achieve a broader overview of the available literature on the topic.

### Research questions

Our research questions were the following: 1) What is known about the relationship between climate change and health, as shown in previous systematic reviews? 2) What are the characteristics of these studies? We registered our plan (CRD42019145972^13^) in PROSPERO, an international prospective register of systematic reviews and followed PRISMA 2020^14^ to report our findings, as a reporting guideline for overviews is still in development.^15^

### Search strategy and selection criteria

To identify relevant studies, we used a systematic search strategy. We included studies in this review if they 1) were systematic reviews of original research and 2) reported at least one health impact as it related (directly or indirectly) to climate change.

We defined a systematic review, based on Cochrane’s definition, as a review of the literature in which one “attempts to identify, appraise and synthesize all the empirical evidence that meets pre-specified eligibility criteria to answer a specific research question [by] us[ing] explicit, systematic methods that are selected with a view aimed at minimizing bias, to produce more reliable findings to inform decision making.”^16^ We included systematic reviews of original research, with or without meta-analyses. We excluded narrative reviews, non-systematic literature reviews and systematic reviews of materials that were not original research (e.g., systematic reviews of guidelines.)

We based our definition of health impacts on the World Health Organization’s (WHO) definition of health as, “a state of complete physical, mental and social well-being and not merely the absence of disease or infirmity.”^17^ Therefore, health impacts included, among others, morbidity, mortality, new conditions, worsening/improving conditions, injuries, and psychological well-being. Climate change (or global warming) could be referred to directly or indirectly, for instance, by synthesizing the direct or indirect health effects of temperature rises or of natural conditions/disasters made more likely by climate change (e.g., floods, wildfires, temperature variability, droughts.) We included systematic reviews whose main focus was not the health impacts of climate change, providing they reported at least one result regarding health effects related to climate change (or consequences of climate change.)

On June 22, 2019, we retrieved systematic reviews regarding the health effects of climate change by searching the electronic databases Medline, CINAHL, Embase, Cochrane, Web of Science using a structured search (see Appendix 1 for final search strategy developed by a librarian.) We did not apply language restrictions. After removing duplicates, we imported references into Covidence.^18^

### Screening process

To select studies, we first screened titles and abstracts to eliminate articles that did not meet our inclusion criteria. Two trained analysts independently screened each article. A senior analyst resolved any conflict or disagreement. Because the topic was new to some team members, to ensure a high-quality screening process, the trained analysts then re-screened all included records and the senior analyst re-screened all excluded records. Two analysts then independently screened the full text of retained articles, again with a senior analyst resolving disagreements.

### Data extraction

Next, we decided on key information that needed to be extracted from studies. We extracted the first author’s name, year of publication, number of studies included, time frame (in years) of the studies included in the article, first author’s institution’s country affiliation, whether the systematic review included a meta-analysis, geographical focus, population focus, the climate impact(s) and the health outcome(s) as well as the main findings and limitations of each systematic review.

Two or more trained analysts (RR, CB, RN, LC, LPB, RAPR) independently extracted data, using Covidence and spreadsheet software (Google Sheets). For analysts who were new to evidence syntheses (CB, LPB, RAPR), the training process included extracting data repeatedly from the same articles to ensure accurate understanding, weekly group meetings to clarify understanding, and daily supervision by more senior team members (RR, RN, HOW). An additional trained analyst from the group or senior research team member resolved disagreements between individual judgments.

### Coding and Data Mapping

To summarize findings from previous reviews, we used a three-step procedure for coding and data mapping. First, to map articles according to climate impacts and health outcomes, two researchers (RR and LC) consulted the titles and abstracts of each article. We developed the categories for climate impacts separately from those for health outcomes and used a mixed approach to coding. We started with an inductive coding method, by identifying categories directly based on our data and followed up with a deductive approach to finalize categories by consulting previous conceptual frameworks of climate impacts and health outcomes.^1,2,19^ The same two researchers independently coded each article according to their climate impact and health outcome. We then compared coding and resolved disagreements through discussion.

Next, still using spreadsheet software, we created a matrix to map articles according to their combination of climate impacts and health outcomes. Each health outcome occupied one row, whereas climate impacts each occupied one column. We placed each article in the matrix according to the combination(s) of their climate impact(s) and health outcome(s). For instance, if we coded an article as ‘extreme weather’ for climate and ‘mental health’ for health impact, we noted it in the cell at the intersection of these two codes. We calculated frequencies for each cell to identify frequent combinations and gaps in literature. Because one study could investigate more than one climate impact and health outcome, the frequency counts for each category could exceed the number of studies included in this review.

Finally, we summarized findings of the studies individually according to their combination of climate impacts and health outcomes. We re-read the Results and Discussion sections of each article as part of this step. We first wrote an individual summary for each study, then we collated the summaries of all studies exploring the same combination of categories to develop an overall summary of findings for each combination of categories.

### Quality assessment

We used a modified version of AMSTAR-2 to assess the quality of the included systematic reviews (Appendix 2). Since AMSTAR-2 was developed for syntheses of systematic reviews of randomized controlled trials, working with a team member with expertise in knowledge synthesis (AT), we adapted it to suit a research context that is not amenable to randomized controlled trials. We used items 5, 6, 10, 11, 12, 14, 15, 16 without modification and modified items 1 to 4, 7 to 9 and 13.

## Results

### Articles identified

As shown in the PRISMA diagram in Figure 1, from an initial set of 2619 references, we retained 94 for inclusion.

**Figure 1.**
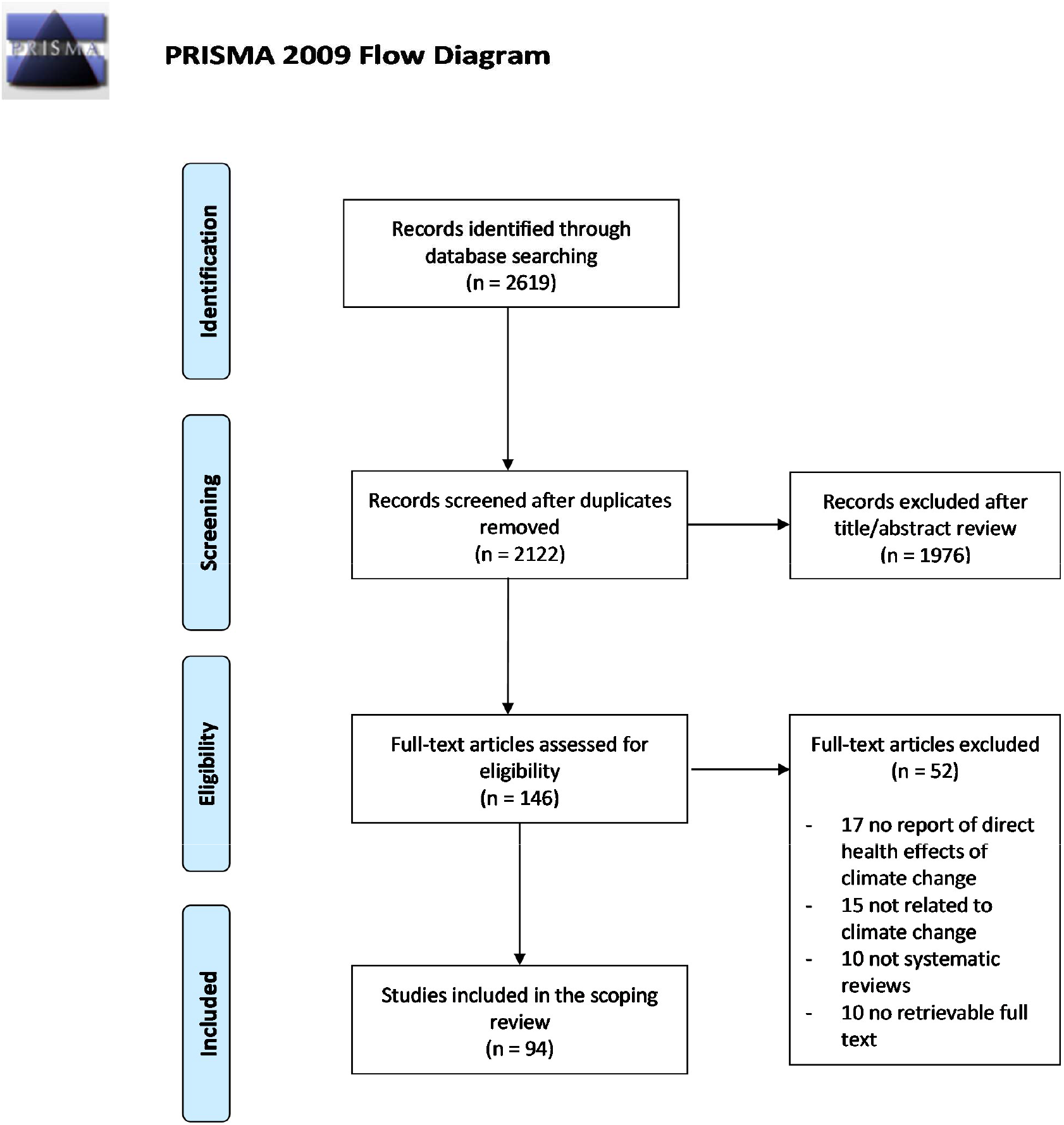
The flow chart for included articles in this review.

### Study Descriptions

A detailed table of all articles and their characteristics can be found in Appendix 3. Publication years ranged from 2007 to 2019 (year of data extraction), with the great majority of included articles (n = 69; 73%) published since 2015 (Figure 2). A median of 30 studies had been included in the systematic reviews (mean = 60; SD = 49; range 7 to 722). Approximately one fifth of the systematic reviews included meta-analyses of their included studies (n = 18; 19%). The majority of included systematic reviews’ first authors had affiliations in high-income countries, with the largest representations by continent in Europe (n = 30) and Australia (n = 24) (Figure 3).

**Figure 2.**
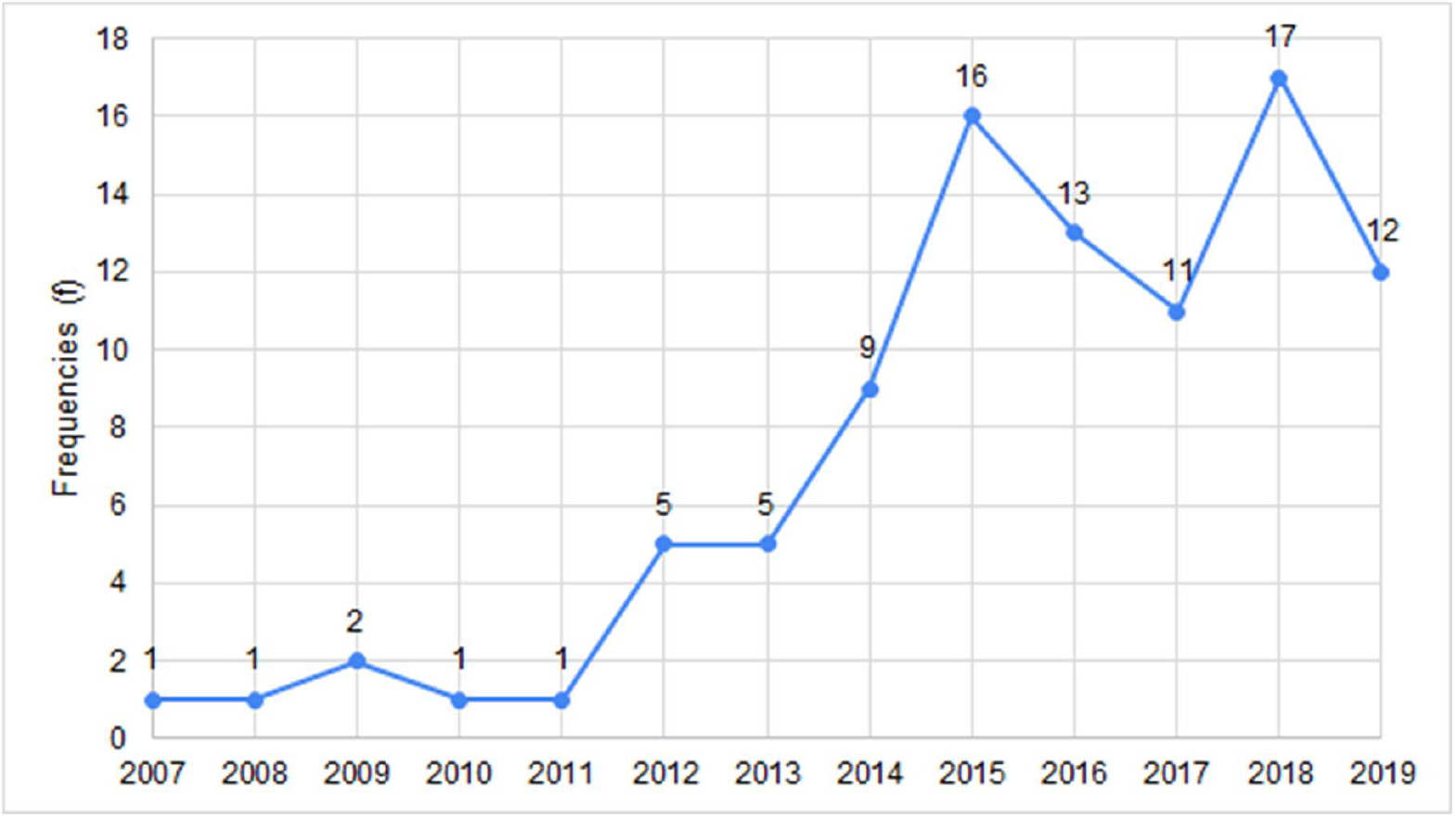
Number of included systematic reviews by year of publication.

**Figure 3.**
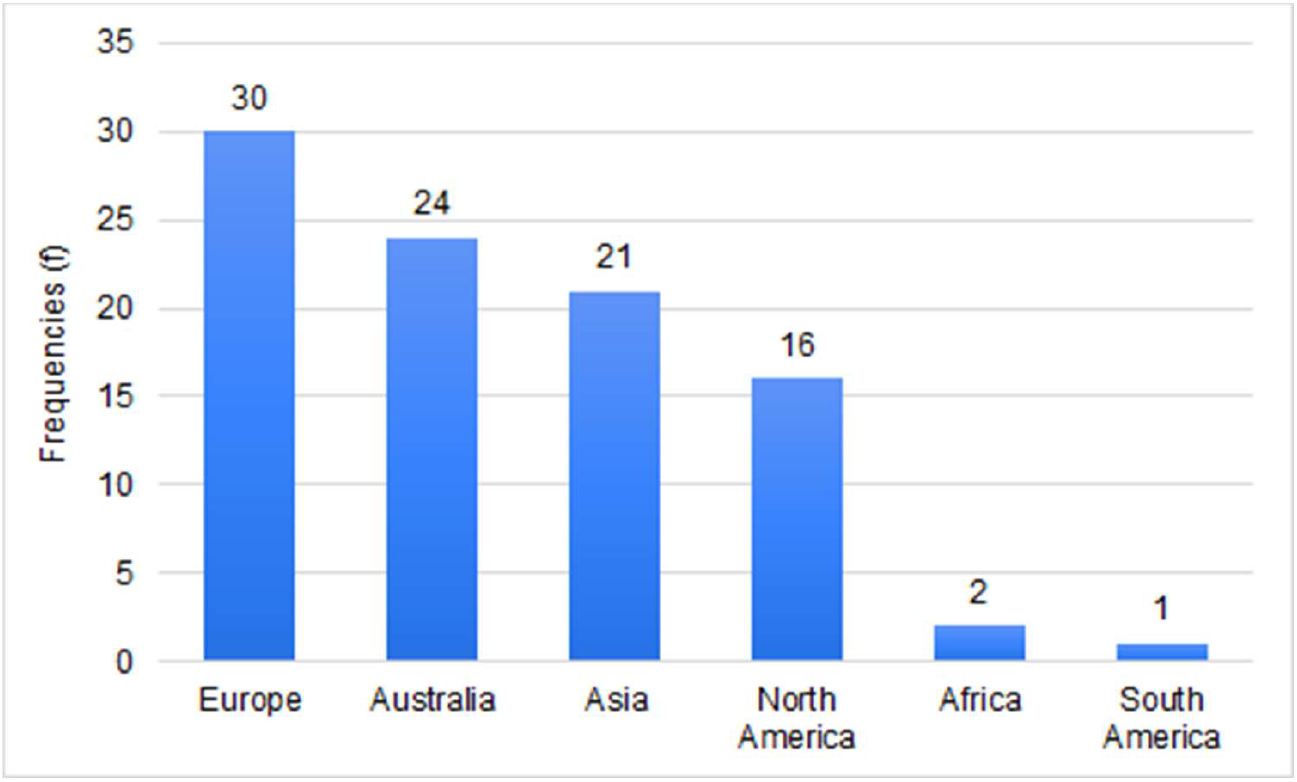
Number of publications according to geographic affiliation of the first author. *Countries of origin within continents in frequency order (highest to lowest frequency) and alphabetical: ***Europe***: United-Kingdom (9), Germany (6), Italy (4), Sweden (4), Denmark (2), France (2), Georgia (1), Greece (1) and Finland (1). ***Australia***: All Australia. ***Asia***: China (11), Iran (4), India (1), Jordan (1), Korea (1), Nepal (1), Philippines (1), Taiwan (1). ***North America:*** United-States (15), Canada (1). ***Africa:*** Ethiopia (1), Ghana (1). ***South America:*** Brazil (1).

Regarding the geographical focus of systematic reviews, most of the included studies (n = 68; 72%) had a global focus or no specified geographical limitations and therefore included studies published anywhere in the world. The remaining systematic reviews either targeted certain countries (n = 12) (1 for each Australia, Germany, Iran, India, Ethiopia, Malaysia, Nepal, New Zealand and 2 reviews focused on China and the United States), continents (n = 5) (3 focused on Europe and 2 on Asia), or regions according to geographical location (n = 6) (1 focused on Sub-Saharan Africa, 1 on Eastern Mediterranean countries, 1 on Tropical countries, and 3 focused on the Arctic), or according to the country’s level of income (n = 3) (2 on low to middle income countries, 1 on high income countries).

Regarding specific populations of interest, most of the systematic reviews did not define a specific population of interest (n = 69; 73%). For the studies that specified a population of interest (n = 25; 26.6%), the most frequent populations were children (n = 7) and workers (n = 6), followed by vulnerable or susceptible populations more generally (n = 4), the elderly (n = 3), pregnant people (n = 2), people with disabilities or chronic illnesses (n = 2) and rural populations (n = 1).

### Quality assessment

We assessed studies for quality according to our revised AMSTAR-2. Out of 94 systematic reviews, the most commonly fully satisfied criterion was #1 (PICO components) with 81/94 (86%) of included systematic reviews fully satisfying this criterion. The next most commonly-satisfied criteria were #16 (potential sources of conflict of interest reported) (78/94 = 83% fully), #13 (account for limitations in individual studies) (70/94 = 75% fully and 2/94 = 2% partially), #7 (explain both inclusion and exclusion criteria) (64/94 = 68% fully and 19/94 = 20% partially), #8 (description of included studies in adequate detail) (36/94 = 38% fully and 41/94 = 44% partially), and #4 (use of a comprehensive literature search strategy) (0/94 = 0% fully and 80/94 = 85% partially). For criteria #11, #12, and #15, which only applied to reviews including meta-analyses, 17/18 (94%) fully satisfied criterion #11 (use of an appropriate methods for statistical combination of results), 12/18 (67%) fully satisfied criterion #12 (assessment of the potential impact of RoB in individual studies) (1/18 = 6% partially), and 11/18 (61%) fully satisfied criterion #15 (an adequate investigation of publication bias, small study bias). Full details are available in Appendix 4.

### Climate Impacts and Health Outcomes

For both the climate impacts and health outcomes, systematic reviews could have a general or a specific focus. A general focus consisted of investigating the general impacts of climate change or multiple impacts simultaneously, whereas a specific focus targeted specifically only one climate impact or health outcome. When combining the climate impact to the health outcome, four combinations became apparent. Table 1 shows these four combinations with sample titles of systematic reviews within that combination. The most frequent combination (n = 52; 55%) consisted of studies investigating a specific climate impact on a specific health outcome (e.g., the impact of floods on mental health) and the least frequent combination (n = 5; 5%) consisted of studies exploring general or multiple climate impacts’ effects on multiple health outcomes (e.g., health impacts of climate change.)

**Table 1.** Summary of the four scenarios possible when combining climate impact and health outcome categories with frequencies and examples of paper titles.

| Frequency (%) and Example Titles |  | Health Outcome |  |
| --- | --- | --- | --- |
|  |  | Multiple (n = 29) | Specific (n = 65) |
| **Climate Impact** | General or multiple (n = 18) | n = 5 (5%)<br>E.g., "Health Impact of Climate Change in Older People: An Integrative Review and Implications for Nursing,"<sup>20</sup> and, "Climate Change and Health in the Eastern Mediterranean Countries: A Systematic Review."<sup>21</sup> | n = 13 (14%)<br>E.g., "Global Warming and Obesity,"<sup>22</sup> and, "Systematic Review of Current Efforts to Quantify the Impacts of Climate Change on Undernutrition."<sup>23</sup> |
|  | Specific (n = 76) | n = 24 (26%)<br>E.g., "Floods and Human Health: A Systematic Review,"<sup>3</sup> and, "Health Effects of Drought: A Systematic Review of the Evidence."<sup>24</sup> | n = 52 (55%)<br>E.g., "The Mental Health Outcomes of Drought: A Systematic Review and Causal Process Diagram,"<sup>25</sup> and, "The Association between Ambient Temperature and Childhood Asthma: A Systematic Review."<sup>26</sup> |

Regarding climate impacts, we identified five mutually exclusive categories, with 13 publications targeting more than one category of climate impacts: 1) Meteorological (n = 71 papers) (e.g., temperature, heat waves, humidity, precipitation), 2) Extreme weather (n = 24) (e.g., water-related, floods, cyclones, hurricanes, drought), 3) Air quality (n = 7) (e.g., air pollution and wildfire smoke exposure), 4) General (n = 5), and 5) Other (n = 3). “General” climate impacts included articles that did not specify climate change impacts but stated general climate change as their focus. “Other” climate impacts included studies investigating other effects indirectly related to climate change (e.g., impact of environmental contaminants) or general environmental risk factors (e.g., environmental hazards, sanitation, and access to clean water.)

We identified ten categories to describe the health outcomes studied by the systematic reviews, and 29 publications targeted more than one category of health outcomes: 1) Infectious diseases (n = 41 papers) (vector-, food-and water-borne), 2) Mortality (n = 32), 3) Respiratory, cardiovascular, cardiopulmonary and neurological (n = 22), 4) Healthcare systems (n = 16), 5) Mental health (n = 13), 6) Pregnancy and birth (n = 11), 7) Dietary (n = 9), 8) Skin and allergies (n = 9), 9) Occupational health and injuries (n = 6) and 10) Other health outcomes (n = 17) (e.g., sleep, arthritis, disability-adjusted life years, non-occupational injuries, etc.)

Figure 4 depicts the combinations of climate impact and health outcome for each study, with Appendix 5 offering further details. The 5 most common combinations are studies investigating the 1) meteorological impacts on infectious diseases (n = 35), 2) mortality (n = 24) and 3) respiratory, cardiovascular, cardiopulmonary and neurological outcomes (n = 17), and 4) extreme weather events’ impacts on infectious diseases (n = 14) and 5) meteorological impacts on health systems (n = 11).

**Figure 4.**
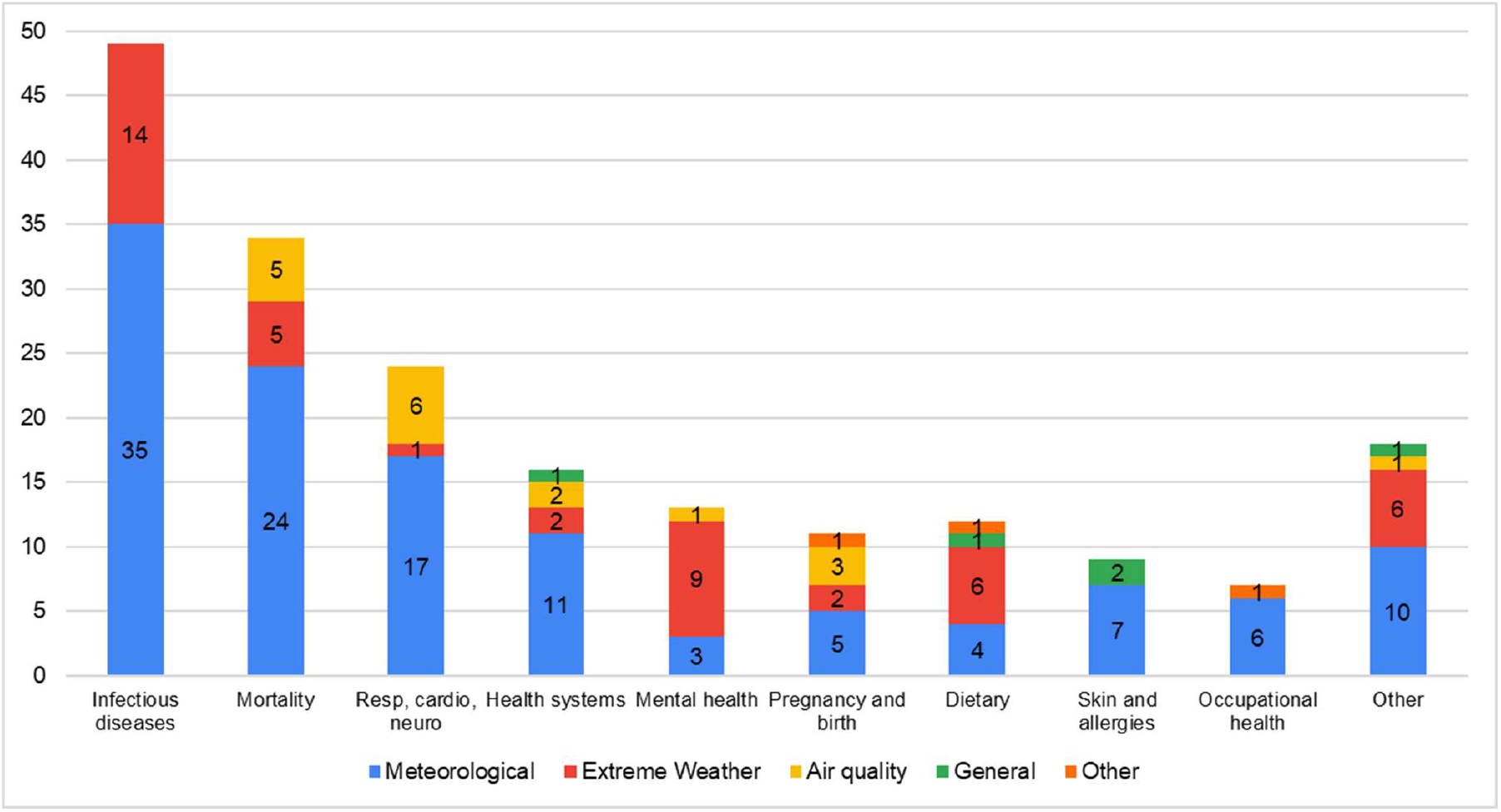
Summary of the combination of climate impact and health outcome (frequencies). *Note:* The total frequency for one category of health outcome could exceed the number of publications included in this health outcome, since one publication could explore the health impact according to more than one climate factor (e.g., one publication could explore both the impact of extreme weather events and temperature on mental health.)

For studies investigating meteorological impacts on health, the three most common health outcomes studied were impacts on 1) infectious diseases (n = 35), 2) mortality (n = 24) and 3) respiratory, cardiovascular, cardiopulmonary and neurological outcomes (n = 17). Extreme weather event studies most commonly reported health outcomes related to 1) infectious diseases (n = 14), 2) mental health outcomes (n = 9) and 3) dietary outcomes (n = 6) and other health outcomes (e.g., injuries, sleep) (n = 6). Studies focused on the impact of air quality were less frequent and explored mostly health outcomes linked to 1) respiratory, cardiovascular, cardiopulmonary and neurological outcomes (n = 6), 2) mortality (n = 5) and 3) pregnancy and birth outcomes (n = 3).

Meteorological factors’ impact on all health outcomes are explored, although some health outcomes are more rarely explored (e.g., mental health and dietary outcomes). In contrast, the impact of extreme weather events and air quality on skin and allergies and occupational health are not explored and their impacts on respiratory, cardiovascular, cardiopulmonary and neurological outcomes, health systems and pregnancy outcomes are only rarely explored. The impacts of air quality on infectious diseases, dietary outcomes, skin and allergies, and occupational health and injuries are also not explored. Most health outcomes are most frequently explored according to the meteorological impacts, however, mental health outcomes and dietary outcomes are most frequently explored according to extreme weather events.

### Summary of Findings

Most reviews suggest a deleterious impact of climate change on multiple adverse health outcomes, with some associations being explored and/or supported with consistent findings more often than others (see Table 2 for a summary of findings according to health outcomes). For instance, the association between meteorological factors, such as temperature and humidity, and vector-borne diseases is quite substantially supported by multiple reviews (n = 22) conducted in multiple geographic locations. In contrast, the association between wildfire smoke exposure and adverse birth outcomes is plausible, but the evidence from included reviews is still in its infancy stage because only a few reviews (n = 3) investigated this association and the findings are currently conflicting.

**Table 2.** Summary of findings from systematic reviews according to health outcome and climate impact. Reviews that covered multiple climate impacts are listed in each relevant category.

| Climate Impact | F | Summary of Findings |
| --- | --- | --- |
| **Infectious diseases (n = 41)** |  |  |
| *Vector borne infectious diseases (n = 25)* |  |  |
| Meteorological | 22 | Systematic reviews suggest that meteorological factors, such as temperature, precipitation, humidity, and wind, are associated with diverse vector-borne infectious diseases, including malaria and dengue.<sup>6,9,20,21,30,32,35–50</sup> This association was mostly proportional (e.g., higher temperature and increased rainfall associated with vector-borne diseases), although findings were at times conflicting, with some suggesting an inversely proportional association<sup>9</sup> (e.g., decreased rainfall) or no association at all<sup>40</sup> (e.g., with the human puumala hantavirus infection.) Geographic location, seasonality and potential interaction with other climate-related factors may partly explain these inconsistencies.<sup>9,30</sup> Temperature, humidity and rainfall were the most common and important meteorological factors reported by reviews and factors such as wind, air pressure and sunshine were reported less often. |
| Extreme weather | 7 | There are limited and conflicting findings concerning the association of extreme weather events with vector-borne diseases. Some reviews suggest water-related extreme events<sup>51</sup> and flooding<sup>3,32,52</sup> are associated with an increased risk of vector-borne diseases, while drought is associated with a reduction of dengue incidence.<sup>9</sup> Other reviews focused specifically on Puerto Rico<sup>43</sup> and Australia<sup>53</sup> did not find an association between hurricanes and/or floods and mosquito-borne disease transmission. |
| *Food and water borne infectious diseases (n = 19)* |  |  |
| Meteorological | 14 | Reviews suggest that meteorological factors, such as temperature, precipitation, and humidity, are associated with diverse food- and water-borne infectious diseases, in particular, cholera, food poisoning, schistosomiasis, salmonella and E. coli.<sup>8,21,32,41,45,48,54–61</sup> Overall, higher temperatures and humidity,<sup>8,41,54,58</sup> along with lower precipitation<sup>21,61</sup> was associated with these infectious diseases (e.g., E. coli<sup>58</sup>; bacterial gastrointestinal infections.<sup>54</sup>) Directionality and strength of the association seemed to vary according to disease and pathogens,<sup>59</sup> seasons, and geographic region.<sup>56</sup> |
| Extreme weather | 10 | Reviews suggest a proportional association between extreme water-related events,<sup>47,51,62</sup> such as flooding<sup>3,41,52</sup> and heavy rainfall<sup>35</sup>, and food- and water-borne diseases, including diarrhea, food contamination, cholera.<sup>3,32,35,41,45,47,51,52,57,62</sup> Drought may also be proportionally associated with food- and water-borne disease,<sup>24,35</sup> but these associations are less consistent than those with water-related extreme events.<sup>57</sup> |
| *Other infectious diseases (n = 8)* |  |  |
| Meteorological | 8 | Reviews suggest an association of most meteorological factors, such as temperature and humidity, with various other infectious diseases, including meningitis,<sup>27,35</sup> Ebola,<sup>27</sup> influenza,<sup>32</sup> and pediatric infectious diseases such as hand-foot-and-mouth disease.<sup>4,5,31,49,55</sup> This association was mostly proportional for meteorological factors such as temperature,<sup>4,5,49</sup> diurnal temperature range,<sup>31</sup> and humidity,<sup>4,5,32</sup> although some meteorological factors, such as air pressure<sup>5</sup> and lower temperatures<sup>32,49</sup> were inversely proportional to these diseases. Some conflicting evidence is reported concerning the association with some meteorological factors, such as sunshine with hand-foot-and-mouth disease,<sup>4,5</sup> and humidity and pediatric infectious diseases.<sup>55</sup> No association was found between some meteorological factors, such as precipitation, wind speed and sunshine with hand-foot-and-mouth disease.<sup>4,5</sup> |
| **Mortality (n = 32)** |  |  |
| Meteorological | 24 | Reviews suggest that temperature (high, low, or diurnal range) was consistently associated with all-cause and cause-specific mortality.<sup>20,21,27,28,31,34,45,47,49,63–76</sup> A strong association was reported between heat (including heat waves) and mortality (all-cause),<sup>63</sup> heat-,<sup>21,68</sup> stroke-,<sup>27,69</sup> cardiovascular-,<sup>34,47</sup> and respiratory-related,<sup>20,34,70</sup> especially in rural,<sup>67</sup> very young children<sup>49</sup> and ageing populations.<sup>28</sup> Mortality seems to be the most frequent health outcome studied in association with heatwaves.<sup>64</sup> Inconsistent results are found concerning the association between heat and childhood mortality.<sup>74</sup> Due to limited evidence, this association was weaker in some geographical regions.<sup>27,71</sup> Also, heat wave intensity (compared to duration) was more strongly associated with heat-related mortality.<sup>75</sup> Finally, although less studied, low temperature was also associated with mortality,<sup>49,76</sup> specifically respiratory,<sup>63</sup> stroke,<sup>69</sup> and cardiovascular mortality.<sup>47,66,70</sup> |
| Extreme Weather | 5 | Reviews suggest an association between extreme weather events such as floods,<sup>3</sup> droughts,<sup>24</sup> cyclones<sup>77</sup> and other water-related events,<sup>20,51</sup> with direct (e.g., drowning) and indirect long-term mortality (e.g., due to malnutrition, environmental toxin exposure, armed conflict, etc.).<sup>3,24,51,77</sup> |
| Air quality | 5 | Reviews suggest an association between exposure to air pollution<sup>20,78</sup> or wildfire smoke<sup>79–81</sup> and air pollution related-mortality, such as respiratory-specific mortality. There is currently limited evidence, but reviews suggest a potential association between wildfire smoke exposure and cardiovascular-specific mortality.<sup>79–81</sup> |
| **Respiratory, neurological, cardiovascular and cardiopulmonary (n = 22)** |  |  |
| Meteorological | 17 | Reviews suggest an association between meteorological factors, such as temperature and humidity, and cardiopulmonary, cardiovascular, respiratory and neurological outcomes.<sup>20,26,27,31,34,37,45,49,55,63,66,68,69,73,74,82,83</sup> Exposure to high temperatures and extreme heat are associated to cardiovascular and respiratory diseases,<sup>20,27,37,49,66</sup> stroke,<sup>69</sup> long-term neurological outcomes (due to heat strokes),<sup>68</sup> myocardial infarction,<sup>34,83</sup> and childhood asthma and pediatric respiratory diseases.<sup>26,74</sup> A review also suggests a beneficial association between heat and the shortening of a respiratory virus season.<sup>45</sup> Exposure to low temperature (cold), temperature drop, or diurnal temperature range was associated with cardiovascular and respiratory diseases,<sup>31,63,66</sup> stroke,<sup>69</sup> and myocardial infarctions.<sup>34</sup> Humidity (most often high humidity, but also lower humidity) and low temperatures were also associated with respiratory diseases in children, including childhood asthma.<sup>26,55,82</sup> |
| Extreme Weather | 1 | A previous review suggests an association between drought and respiratory, cardiovascular and cardiopulmonary outcomes, most likely due to droughts leading to increased dust in the air.<sup>24</sup> |
| Air quality | 6 | Reviews suggest a proportional association between exposure to air pollution<sup>20,21,45</sup> or wildfire smoke exposure<sup>79–81</sup> and respiratory outcomes, including asthma, chronic obstructive pulmonary disease, coughing, wheezing, and overall lung function. Although there is currently limited evidence,<sup>79</sup> reviews also suggest a potential association between air pollution or wildfire smoke exposure and cardiovascular outcomes.<sup>45,80,81</sup> |
| **Health systems (n = 16)** |  |  |
| General | 1 | A previous review suggests that climate change in general puts a strain on public health resources, via population health issues and shows that using an integrated surveillance system may guide future adaptation to climate change.<sup>84</sup> |
| Meteorological | 11 | Previous reviews suggest an association between temperature change<sup>31</sup> extreme heat, aridity and cold temperatures and an increase in use of healthcare services (mostly linked to heat-related health impacts), such as an increase in emergency department visits, hospital admissions and use of ambulances.<sup>20,21,27,31,34,49,64,71,74,83,85</sup> |
| Extreme weather | 2 | Reviews suggest that extreme weather events<sup>33</sup> and flooding<sup>3</sup> are associated with an increase in use of healthcare services (e.g., increased hospitalizations) and a compromised quality of care as extreme weather events may lead to power outages.<sup>33</sup> |
| Air quality | 2 | Reviews suggest an association between wildfire smoke exposure and an increase in use of healthcare services, such as an increase in emergency department visits.<sup>79,81</sup> |
| **Mental health (n = 13)** |  |  |
| Meteorological | 3 | Reviews suggest an association of most meteorological factors such as temperature increase, aridity, heat, and heat waves with mental health outcomes, including hospital admissions for mental health reasons,<sup>21</sup> suicide,<sup>86</sup> and exacerbation of pre-existing mental health conditions, difficulty sleeping, and fatigue.<sup>83</sup> No association was found between sunlight duration and suicide incidence.<sup>86</sup> |
| Extreme weather | 9 | Most reviews reported a proportional association of extreme weather events,<sup>45,51,87,88</sup> flooding,<sup>3,20,89</sup> and drought<sup>24,25</sup> with diverse mental health issues, including, psychological distress, post-traumatic stress disorder, anxiety, depression, psychotropic medication use, alcohol consumption. There was conflicting evidence regarding the association of floods with suicide, tobacco, alcohol and substance abuse.<sup>89</sup> No association was found between drought and suicide.<sup>24</sup> |
| Air quality | 1 | A previous review suggests no association between wildfire smoke exposure and mental health, as measured by physician visits and hospitalizations for mental health reasons during wildfires.<sup>80</sup> |
| **Pregnancy and birth outcomes (n = 11)** |  |  |
| Meteorological | 5 | Reviews suggest that adverse birth outcomes are higher among people exposed to meteorological factors such as high temperature, heat, sunlight intensity, cold and humidity.<sup>21,90–93</sup> These outcomes include low birth weight, preterm birth, eclampsia and preeclampsia, hypertension and length of pregnancy.<sup>21,90–93</sup> The association between heat and adverse birth outcomes seems to have stronger support than the association with cold temperatures.<sup>93</sup> |
| Extreme Weather | 2 | Reviews suggest an association of extreme weather events<sup>87</sup> and flooding<sup>3</sup> with adverse birth outcomes, such as low birth weight, preterm birth and pre-eclampsia. It is suggested that extreme weather events may indirectly affect birth outcomes via the pregnant person's well-being (e.g., stress and worry during pregnancy.)<sup>3,87</sup> |
| Air quality | 3 | There is limited and inconsistent evidence concerning the association between wildfire smoke exposure and adverse birth outcomes, but reviews suggest a potential proportional association between wildfire smoke exposure and lower birth weight.<sup>79–81</sup> |
| Other | 1 | The association between environmental pollutants and adverse birth outcomes (i.e., preterm birth) remains unclear due to conflicting evidence.<sup>29</sup> |
| **Dietary (n = 9)** |  |  |
| General | 1 | A review suggests an association between climate change and obesity.<sup>22</sup> |
| Meteorological | 4 | Reviews suggest an association between meteorological factors, such as changes in temperature, heat and precipitation, with diverse dietary outcomes, including undernutrition, malnutrition and child stunting.<sup>21,23,27,71</sup> This association may be |
|  |  | explained by the impact of meteorological factors, such as temperature increase and precipitation decrease, on crop production and food insecurity.<sup>21,71</sup> |
| Extreme Weather | 6 | Reviews suggest an association between extreme weather events, such as flooding and droughts,<sup>24</sup> and diverse dietary outcomes, including malnutrition and undernutrition in children and adults<sup>21,23,35,45,47</sup> via, amongst others, crops production and food insecurity (e.g., low food aid following flooding<sup>21</sup>). |
| Other | 1 | A review suggests an association between certain environmental risk factors (e.g., sanitation, cooking fuels and food-borne mycotoxins), and childhood stunting, which could be aggravated by climate change.<sup>94</sup> |
| **Skin and allergies (n = 9)** |  |  |
| General | 2 | Reviews suggest a proportional association between climate change, in general, and skin and soft tissue infections (e.g., fatal vibrio vulnificus necrotizing)<sup>95</sup> and ragweed pollen allergies in Europe.<sup>96</sup> |
| Meteorological | 7 | Reviews suggest an association of meteorological factors, such as ultraviolet light exposure, temperature and humidity, with diverse skin and allergic diseases, including skin cancer, sunburn, acute urticaria, eczema and pediatric allergies and skin irritabilities.<sup>27,45,47,49,55,83,97</sup> Higher temperature and ultraviolet light exposure is proportionally associated with sunburn<sup>83</sup> and skin cancer,<sup>45,97</sup> while low humidity and low temperatures were associated with eczema and skin irritabilities in children.<sup>49,55</sup> |
| **Occupational health and injuries (n = 6)** |  |  |
| Meteorological | 6 | Reviews suggest that heat is associated with adverse occupational health outcomes, including injuries, heat strain, dehydration and kidney diseases.<sup>98–103</sup> The most frequent injuries consist of 'slips, trips, falls, wounds, lacerations and amputations.'<sup>99</sup> This association was found in many occupational settings, including agriculture, construction, transport and fishing, and seems to affect both outdoor and indoor workers.<sup>98</sup> This association may be explained by a combination of direct (e.g., dehydration) and indirect factors (e.g., impaired cognitive and physical performance.)<sup>102</sup> |
| Other | 1 | A review suggests an association between environmental pollution (e.g., heavy metals, fertilizers, etc.) and occupational diseases, such as chronic kidney disease.<sup>103</sup> This association is suggested to be affected by increasing temperatures. |
| **Other (n = 17)** |  |  |
| General | 1 | A review suggests an association between climate change in general and disability-adjusted life years, which is an indicator that quantifies 'the burden of disease attributable to climate change'.<sup>104</sup> Authors suggest that the cost of disability-adjusted life years could be high, especially in low to middle income countries. |
| Meteorological | 10 | Reviews suggests an association between increasing temperatures and temperature changes,<sup>31</sup> and other various health outcomes, including acute gouty arthritis,<sup>105</sup> unintentional injuries,<sup>106</sup> diabetes,<sup>63</sup> genitourinary diseases,<sup>31,63</sup> impaired sleep time and quality,<sup>107</sup> cataracts (indirectly associated via people spending more time outside and therefore increased exposure to ultraviolet light),<sup>45,47</sup> heat stress, heat exhaustion and kidney failure,<sup>83</sup> and renal diseases, fever and electrolyte imbalance in children.<sup>49,74</sup> |
| Extreme weather | 6 | Reviews suggests an association between extreme weather events,<sup>88</sup> such as flooding,<sup>3</sup> cyclones,<sup>77</sup> hurricanes,<sup>107</sup> and drought,<sup>24</sup> and other various health outcomes including injuries (e.g., debris, diving in water that is shallower than expected),<sup>3,24,77,88</sup> impaired sleep,<sup>107</sup> esophageal cancer (likely linked to high salinity of water due to droughts),<sup>24</sup> and exacerbation of chronic illnesses.<sup>3,87</sup> |
| Air quality | 1 | There is limited evidence, but a systematic review suggests an association between wildfire smoke exposure and ophthalmic outcomes, such as eye irritation and cataracts.<sup>79</sup> |

Most reviews concluded by calling for more research, noting the limitations observed among the studies included in their reviews, as well as limitations in their reviews themselves. These limitations included, amongst others, some systematic reviews having a small number of publications,^27,28^ language restrictions such as including only papers in English,^20,23^ arriving at conflicting evidence,^29^ difficulty concluding a strong association due to the heterogeneity in methods and measurements or the limited equipment and access to quality data in certain contexts,^27,30–32^ and most studies included were conducted in high-income countries.^33,34^

Previous authors also discussed the important challenge related to exploring the relationship between climate change and health. Not only is it difficult to explore the potential causal relationship between climate change and health, mostly due to methodological challenges, but there are also a wide variety of complex causal factors that may interact to determine health outcomes. Therefore, the possible causal mechanisms underlying these associations were at times still unknown or uncertain and the impacts of some climate factors were different according to geographical location and specificities of the context. Nonetheless, some reviews offered potential explanations for the climate-health association, with the climate factor at times, having a direct impact on health (e.g., flooding causing injuries) and in other cases, having an indirect impact (e.g., flooding causing stress which in turn may cause adverse birth outcomes.)

## Discussion

### Principal results

In this overview of systematic reviews, we aimed to develop an overview of systematic reviews of health impacts of climate change by mapping the characteristics and findings of studies exploring the relationship between climate change and health. We identified four key findings.

First, the most common climate impact studied by included publications consists of meteorological impacts (e.g., temperature, heat, precipitation and humidity), which aligns with findings from a previous scoping review on the health impacts of climate change in the Philippines.^7^ Although this may not be surprising given that a key implication of climate change is the rise in temperature, this finding suggests we also need to undertake research focused on other climate impacts on health, such as the impact of droughts and wildfire smoke, to better prepare for the health crises that arise from these multiple climate-related impacts.

Second, systematic reviews primarily focus on physical health outcomes, such as infectious diseases, mortality, and respiratory, cardiopulmonary, cardiovascular and neurological outcomes, which also aligns with the country-specific previous scoping review.^7^ Regarding mortality, we support Campbell and colleagues’^64^ suggestion that we should expand our focus to include other types of health outcomes. This will allow us to better mitigate and adapt to the full range of threats of climate change.

It is unclear whether the distribution of frequencies of health outcomes reflects the actual burden of health impacts of climate change, or if the most frequently reported outcomes reflect a bias of Western definitions of health. The most commonly-studied health outcomes do not necessarily reflect the definition of health presented by the WHO as, “a state of complete physical, mental and social well-being and not merely the absence of disease or infirmity.”^17^ This suggests that future studies should investigate in greater depth the impacts of climate change on mental and broader social well-being. Indeed, some reviews suggested that climate change impacts psychological and social well-being, via broader consequences, such as political instability, health system capacity, migration, and crime,^83,87^ thus illustrating how our personal health is determined not only by biological and environmental factors but also by social and health systems.

Interestingly, the reviews that explored the mental health impacts of climate change were focused mostly on the direct impacts of experiencing extreme weather events. However, psychologists are also warning about indirect mental health impacts of climate change, which are becoming more prevalent for children and adults alike.^108,109^ Even people who do not experience direct climate impacts, such as extreme weather events, report experiencing disruptive negative emotions when thinking of the destruction of our environment or when worrying about one’s uncertain future and the lack of actions being taken. To foster emotional resilience in the face of climate change, these mental health impacts of climate change need to be further explored. Humanity’s ability to adapt to and mitigate climate change ultimately depends on our emotional capacity to face this threat.

Third, there is a notable geographic difference in the country affiliations of first authors, with three quarters of systematic reviews having been led by first authors affiliated to institutions in Europe, Australia, or North America. While perhaps unsurprising given the inequalities in research funding and institutions concentrated in Western countries, this is of critical importance given the significant health impacts that will be faced in other parts of the world. Research funding organizations should seek to provide more resources to authors in low- to middle-income countries to ensure their expertise and perspectives are better represented in the literature.

Fourth, overall, most reviews suggest an association between climate change and the deterioration of health in various ways, thus illustrating the interdependence of our health and well-being with the well-being of our environment. At times, climate change and its related environmental events may impact health directly (e.g., heat’s impact on dehydration and exhaustion) and other times, it may impact it indirectly (e.g., via behaviour change due to heat.) The climate-health link has been the target of more research in recent years and it is also receiving increasing attention in both public health and climate communication literature.^110,111^ The health framing of climate communication also has implications for healthcare professionals^112^ and policymakers, as these actors could play a key part in climate communication, adaptation, and mitigation. These key stakeholders’ perspectives on the climate-health link, as well as their perceived role in climate adaptation and mitigation could be explored,^113^ since research suggests that health professionals are important voices in climate communications^112^ and especially since, ultimately, these adverse health outcomes will engender pressure on and cost to our health systems and health workers.

### Strengths and Limitations

To the best of our knowledge, the current study provides the first broad overview of previous systematic reviews exploring the health impacts of climate change. Our review has three main strengths. First, by targeting systematic reviews, we achieve a higher-order summary of findings than what would have been possible by consulting individual original studies. Second, by synthesizing findings across all included studies and according to the combination of climate impact and health outcome, we offer a clear, detailed, and unique summary of the current state of evidence and knowledge gaps about how climate change may influence human health. This summary may be of use to researchers, policymakers, and communities. Third, we included studies published in all languages about any climate impact and any health outcome. In doing so, we provide a comprehensive and robust overview.

Our work has three main limitations. First, we were unable to access some full texts and therefore some studies were excluded, even though we deemed them potentially relevant after title and abstract inspection. Other potentially relevant systematic reviews may be missing due to unseen flaws in our systematic search. Second, due to the heterogeneity of the included systematic reviews and the relatively small proportion of studies reporting meta-analytic findings, we could not conduct meta-meta-analyses of findings across reviews. Future research is needed to quantify the climate and health links described in this review, as well as to investigate the causal relationship and other interacting factors. Third, due to limited resources, we did not assess overlap between the included reviews concerning the studies they included. Frequencies and findings should be interpreted with potential overlap in mind.

## Conclusions

Overall, systematic reviews of the health impacts of climate change suggest an association between climate change and the deterioration of health in multiple ways, generally in the direction that climate change is associated with adverse human health outcomes. This is worrisome since these outcomes are predicted to rise in the near future, due to the temperature rise and increase in climate-change-related events such as extreme weather events and worsened air quality. Most studies included in this review focused on meteorological impacts of climate change on adverse physical health outcomes. Future studies could fill knowledge gaps by exploring other climate-related impacts and broader psychosocial health outcomes. Moreover, studies on health impacts of climate change have mostly been conducted by first authors affiliated with institutions in high-income countries. This inequity needs to be addressed, considering that the impacts of climate change are and will continue to predominantly impact lower-income countries. Finally, although most reviews also recommend more research to better understand and quantify these associations, to adapt to and mitigate climate change’s impacts on health, it will also be important to unpack the ‘what, how, and where’ of these effects. Health effects of climate change are unlikely to be distributed equally or randomly through populations. It will be important to mitigate the changing climate’s potential to exacerbate health inequities.

## Data Availability

Data consists of previously published systematic reviews, which are available to readers.

## Acknowledgements

The authors gratefully acknowledge the contributions of Selma Chipenda Dansokho, as research associate, and Thierry Provencher, as research assistant, to this project, and of Frederic Bergeron, for assistance with search strategy, screening and selection of articles for the systematic review.

## Authors’ Contributions

RN, CF, ACT, HOW contributed to the design of the study. CB, RN, LPB, RAPR and HOW contributed to the systematic search of the literature and selection of studies. RR, HOW, LC conducted data analysis and interpretation. RR and HOW drafted the first version of the article with early revision by CB, LC and RN. All authors critically revised the article and approved the final version for submission for publication. RR and HOW had full access to all the data in the study and had final responsibility for the decision to submit for publication.

## APPENDICES

## Appendix 1. Search Strategy

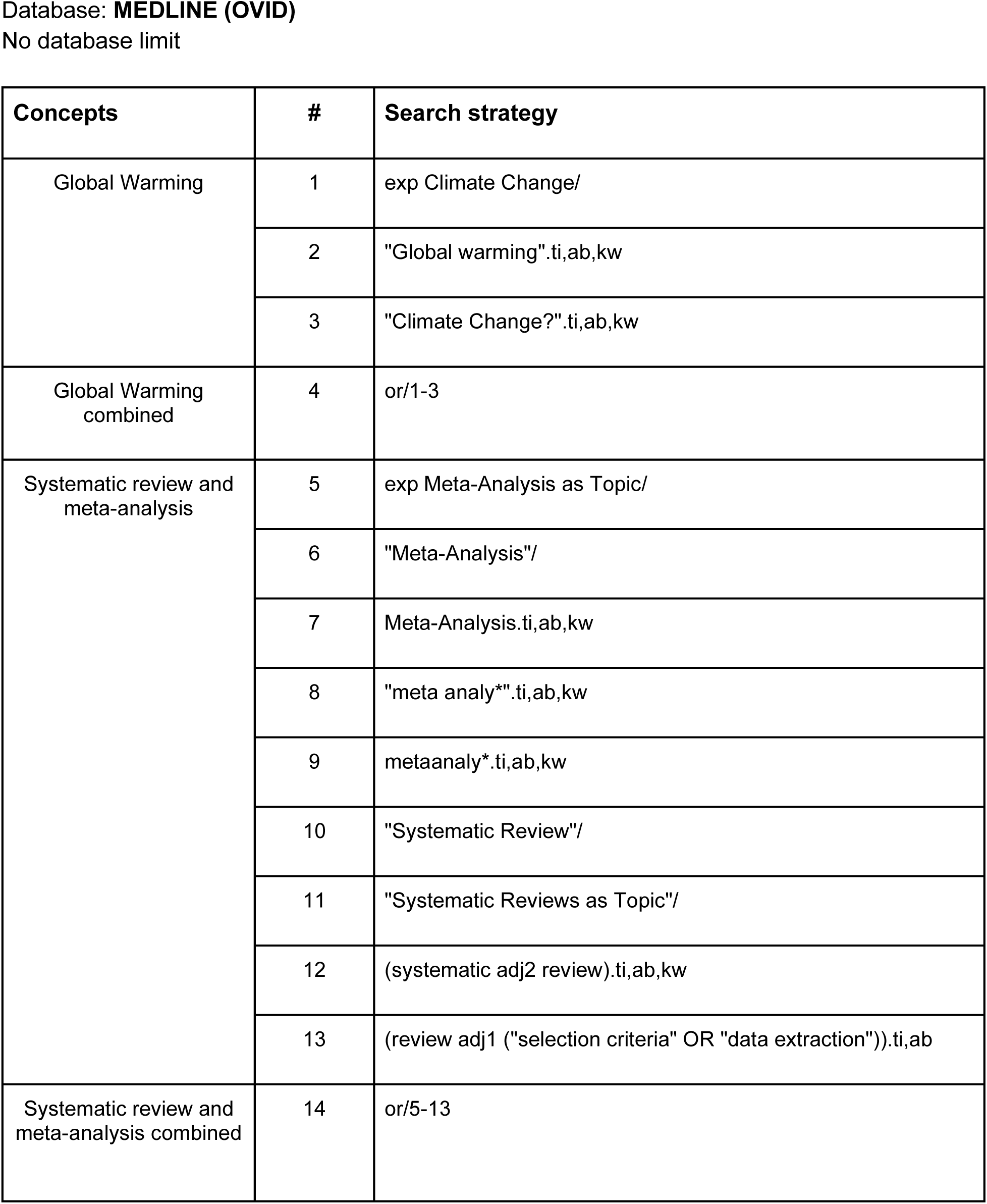
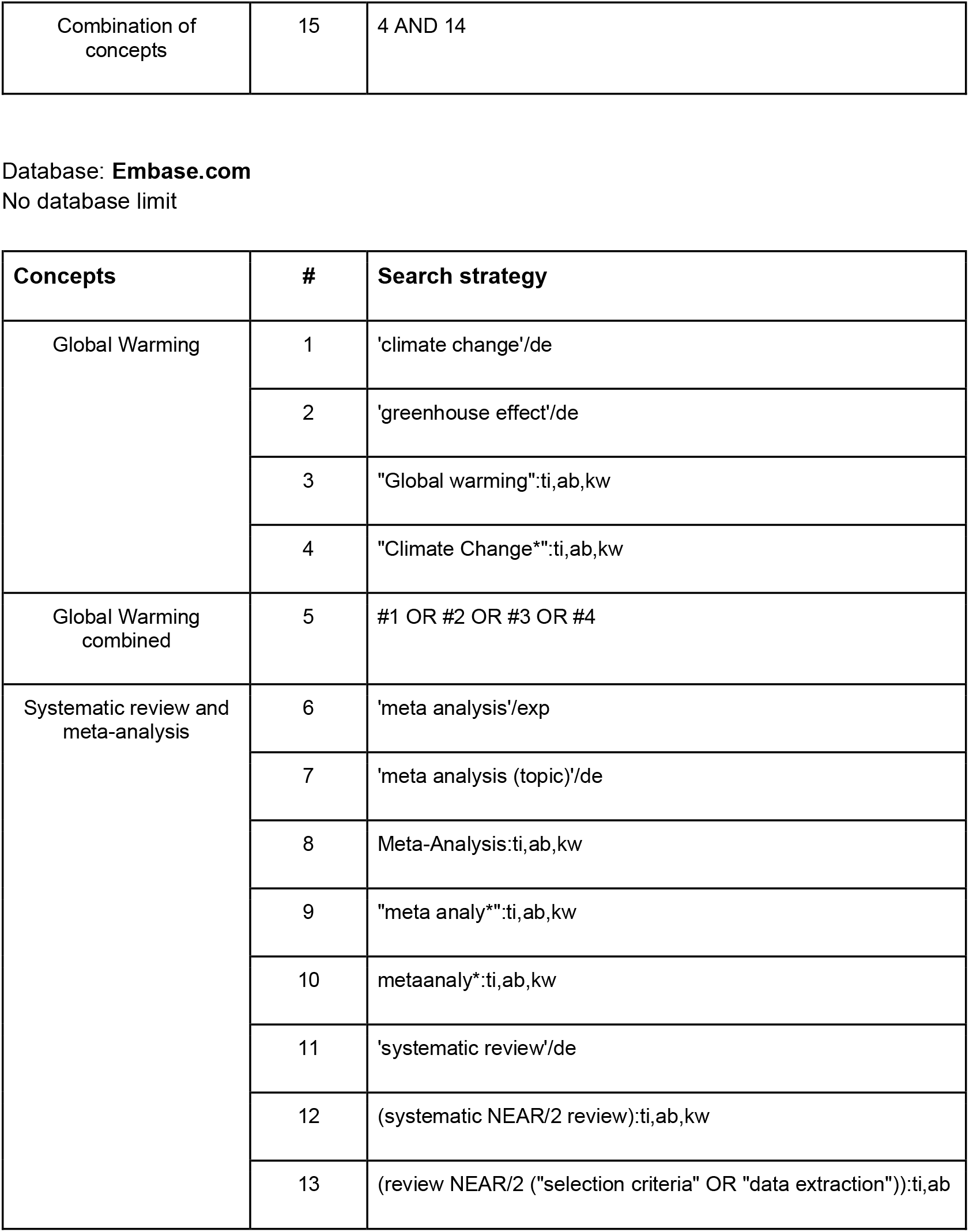
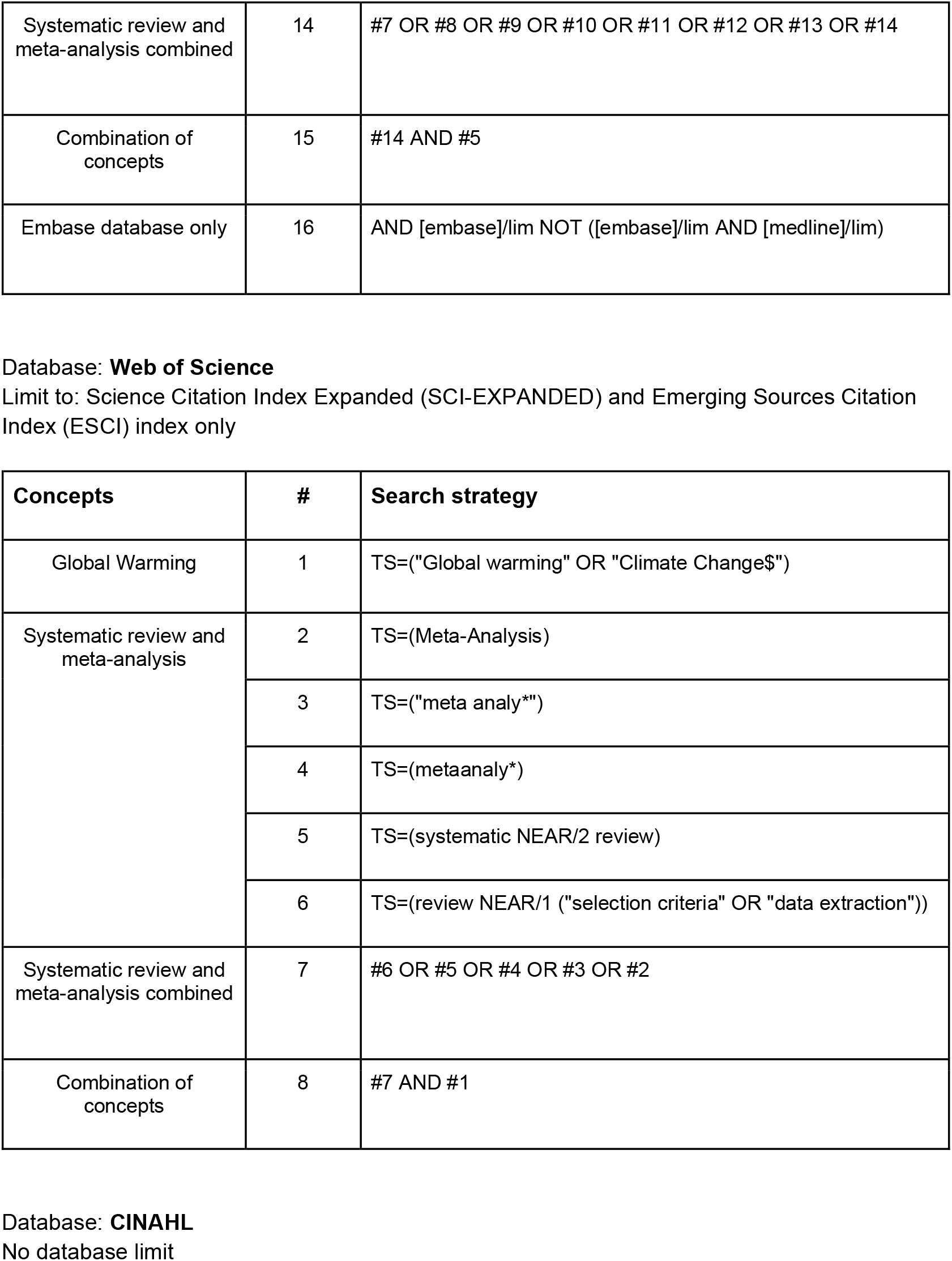
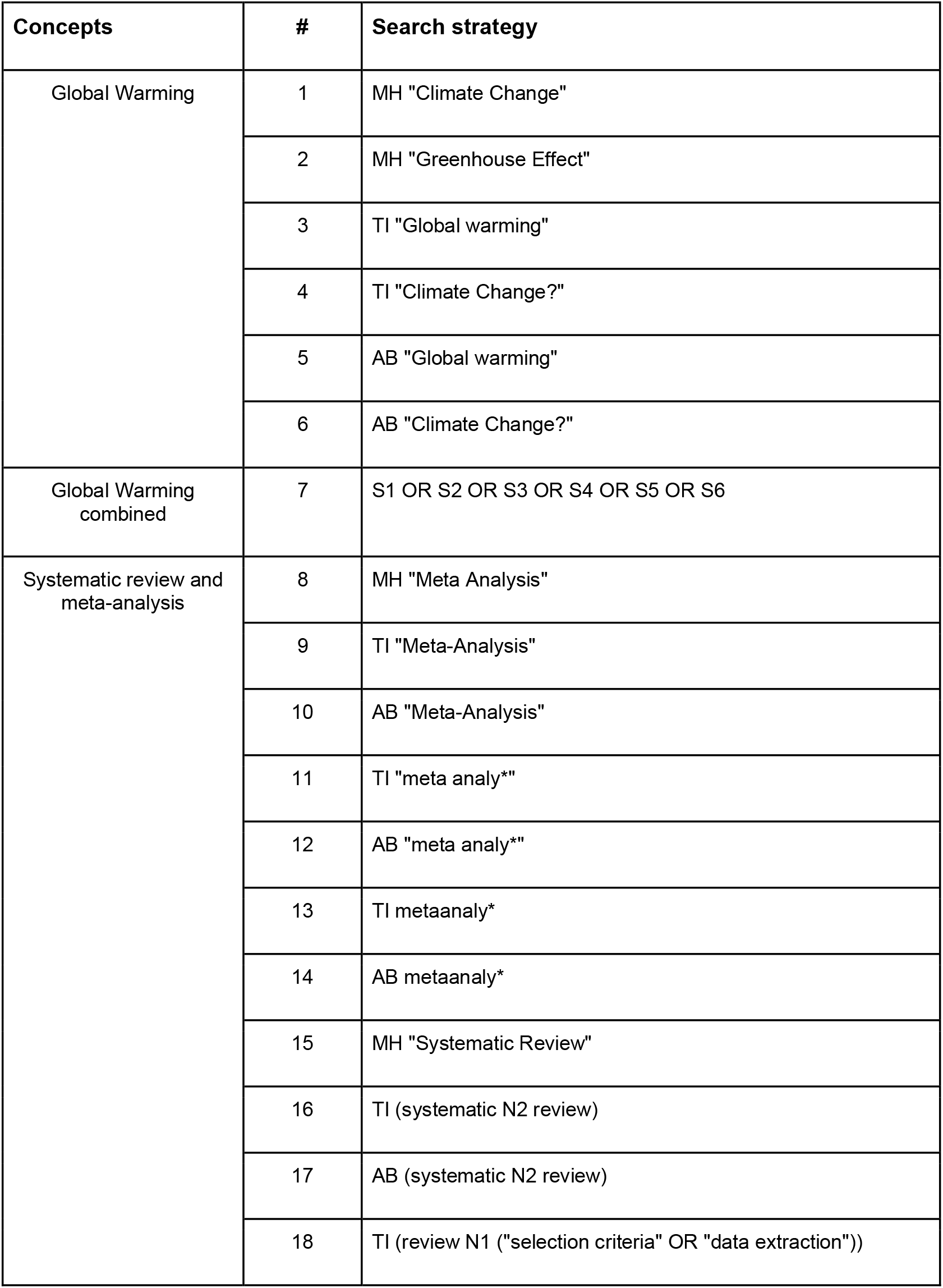
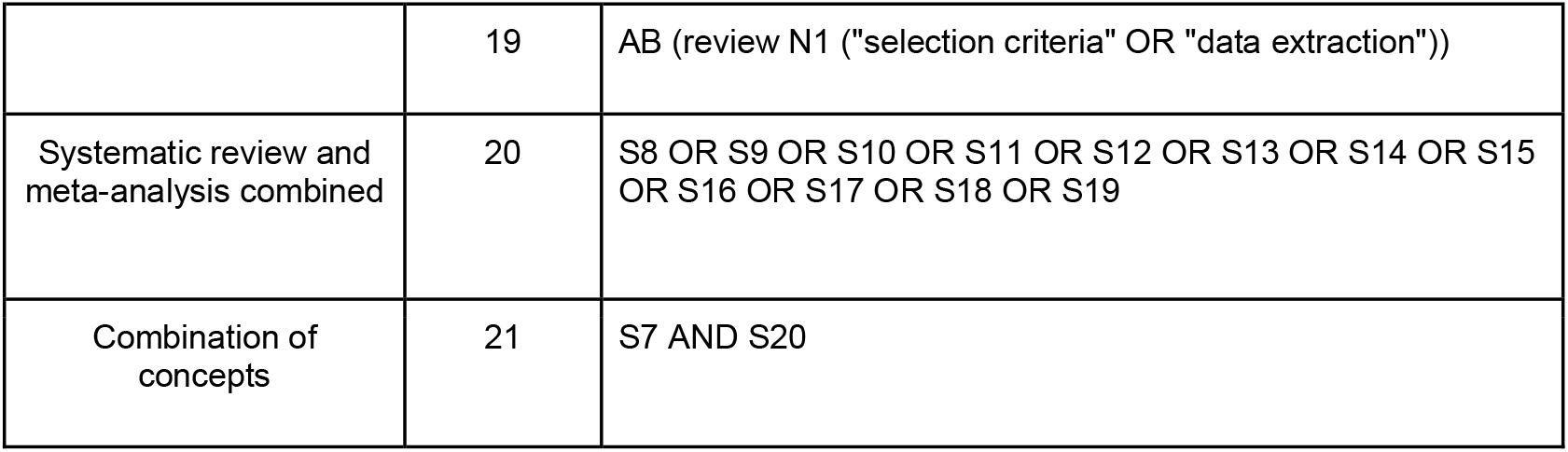

## Appendix 2. Summary of AMSTAR-2 items and modified AMSTAR-2 items.

| Item # | AMSTAR-2 Original Item | AMSTAR-2 Modifications |
| --- | --- | --- |
| 1 | Did the research questions and inclusion criteria for the review include components of PICO?<br>- Population<br>- Intervention<br>- Comparator group<br>- Outcome<br>- Timeframe for follow-up (optional) | “Population” became “Population and/or location”.<br>“Intervention” became “Exposure”.<br>The “Comparator group” category was taken out.<br>A new section (#1.b)) was created, it includes “Definition of the exposure”, “Definition of the outcome” and “Timeframe for follow up”. |
| 2 | Did the report of the review contain an explicit statement that the review methods were established prior to the conduct of the review and did the report justify any significant deviations from the protocol? | To score “yes”, a protocol must have been established before the review. There are no sub-criterias, you can only score yes or no. |
| 3 | Did the review authors explain their selection of the study designs for inclusion in the review? | If the study designs are specified, you score “partial yes”. They must be explained to score “yes”. No specific study design is required. |
| 4 | Did the review authors use a comprehensive literature search strategy? | The “searched trial/study registries” category was taken out.<br>Justified publication restrictions (e.g. language) moved from (partial yes) to (yes) |
| 5 | Did the review authors perform study selection in duplicate? | No modifications. |
| 6 | Did the review authors perform data extraction in duplicate? | No modifications. |
| 7 | Did the review authors provide a list of excluded studies and justify the exclusion? | The explanation of the inclusion and exclusion criteria is evaluated.<br>If there is only one out of the two, you score “partial yes”. The two must be explained to score “yes”. |
| 8 | Did the review authors describe the included studies in adequate detail? | “Populations” became “Populations and/or locations”.<br>“Interventions” became “Exposures”.<br>“Comparator groups” became “Comparator groups (if applicable)”.<br>“Populations and/or locations”, “Exposures” and “Outcomes” must be described in details to score “yes” |
| 9 | Did the review authors use a satisfactory technique for assessing the risk of bias (RoB) in individual studies that were included in the review? | “RoB” became “limitations”. Instead of assessing the RoB, the review authors must have used a satisfactory technique for assessing the limitations in individual studies that were included in the review. |
| 10 | Did the review authors report on the sources of funding for the studies included in the review? | No modifications. |
| 11 | If meta-analysis was performed did the review authors use appropriate methods for statistical combination of results? | No modifications. |
| 12 | If meta-analysis was performed, did the review authors assess the potential impact of RoBin individual studies on the results of the meta-analysis or other evidence synthesis | No modifications. |
| 13 | Did the review authors account for RoB in individual studies when interpreting/discussing the results of the review? | “RoB” became “limitations”. Instead of accounting for RoB in individual studies, the review authors must have accounted for limitations when interpreting/ discussing the results of the review. |
| 14 | Did the review authors provide a satisfactory explanation for, and discussion of, any heterogeneity observed in the results of the review? | No modifications. |
| 15 | If they performed quantitative synthesis did the review authors carry out an adequate investigation of publication bias (small study bias) and discuss its likely impact on the results of the review? | No modifications. |
| 16 | Did the review authors report any potential sources of conflict of interest, including any funding they received for conducting the review? | No modifications. |

## Appendix 3. Overview of included studies (N=94) according to study characteristics and summary of climate impacts on health outcomes. Studies are presented in alphabetical order according to the first author’s last name.

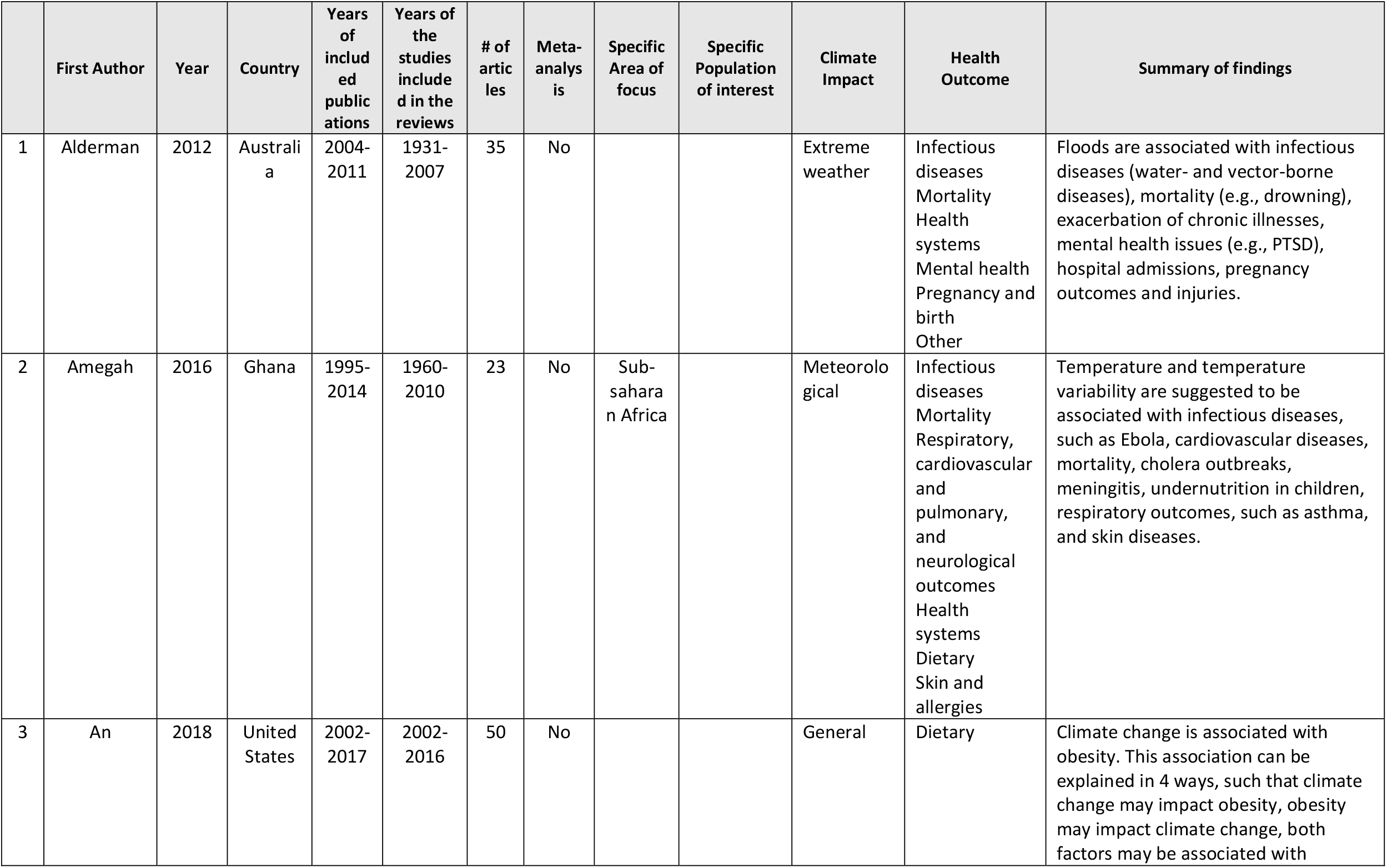
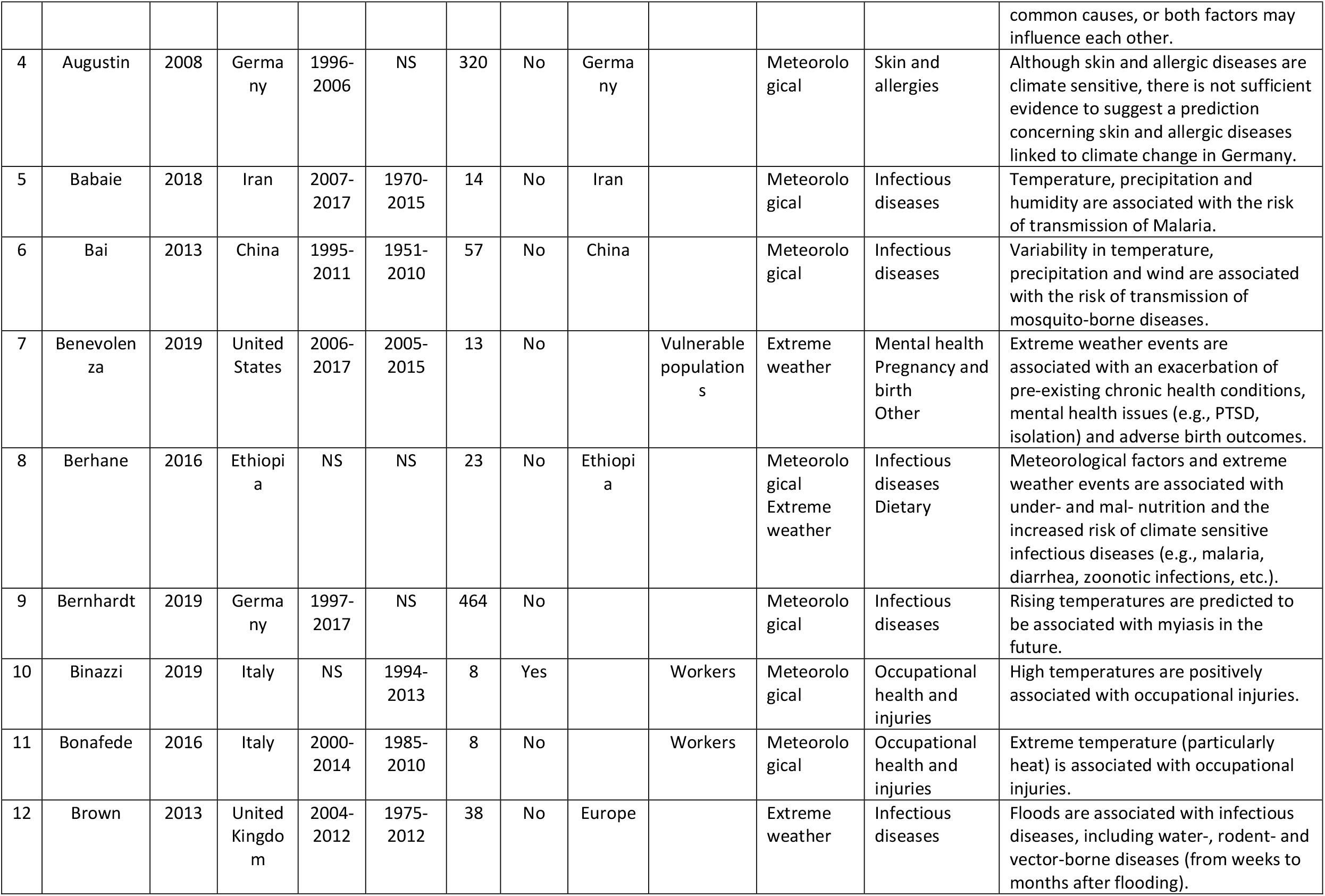
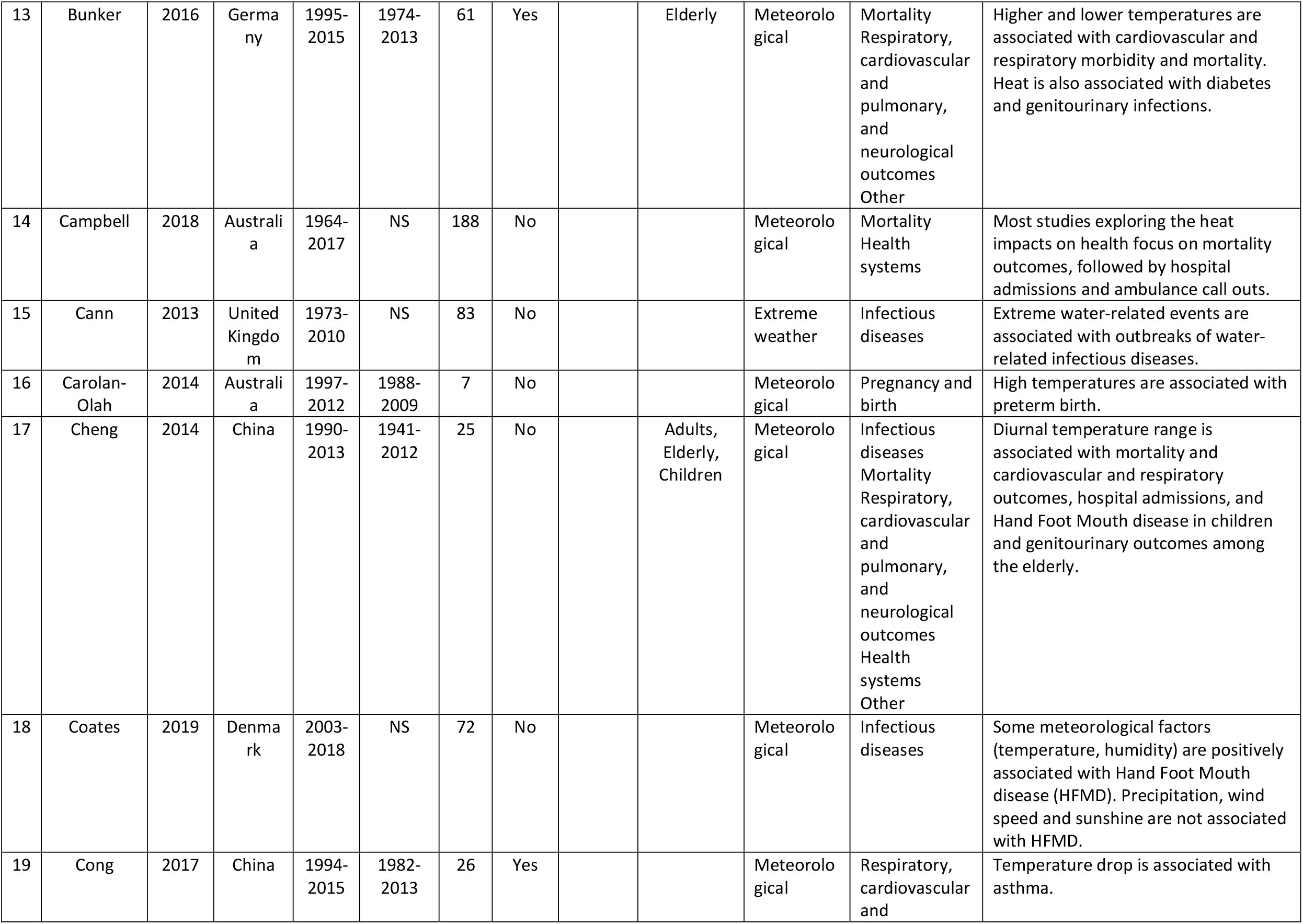
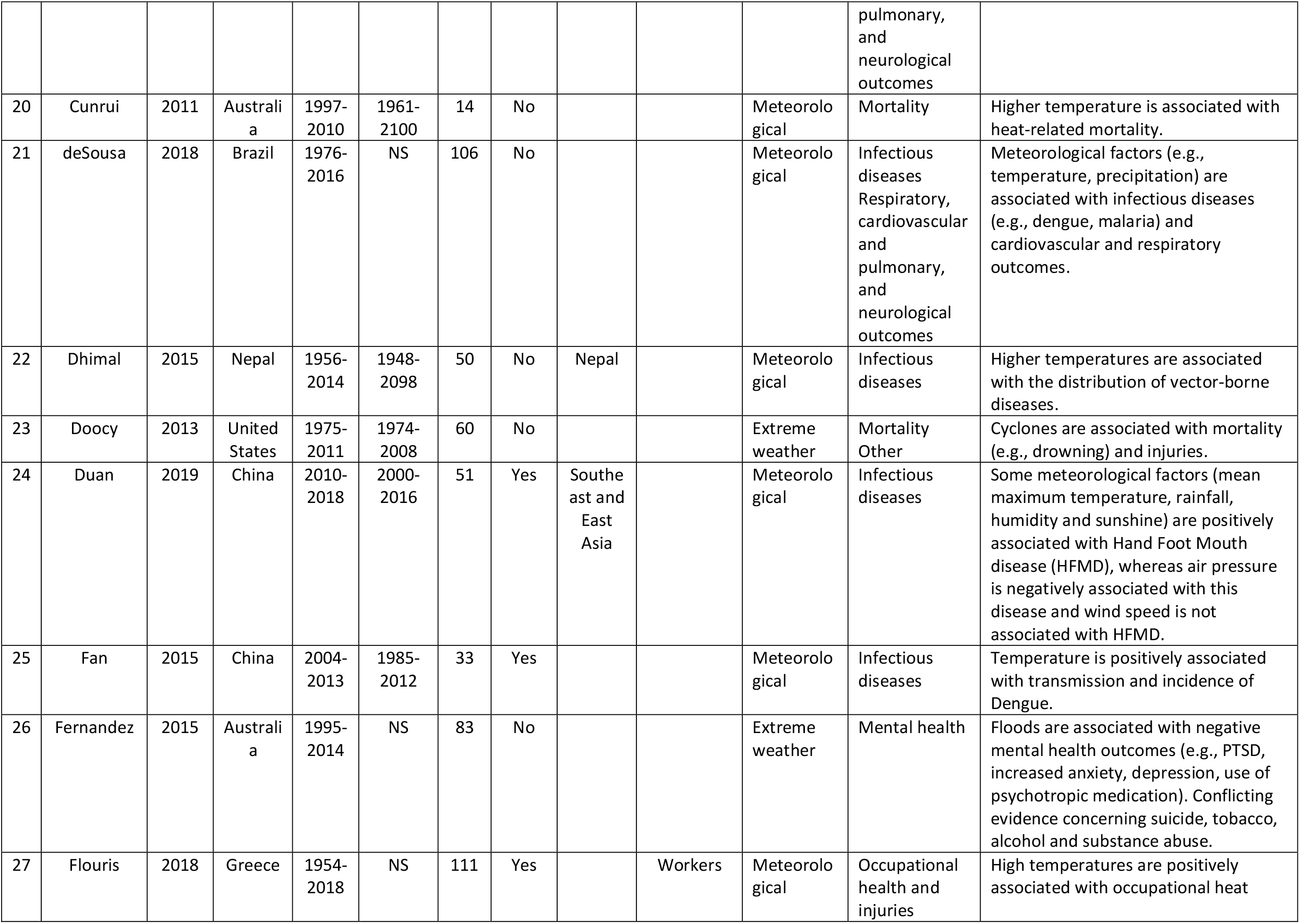
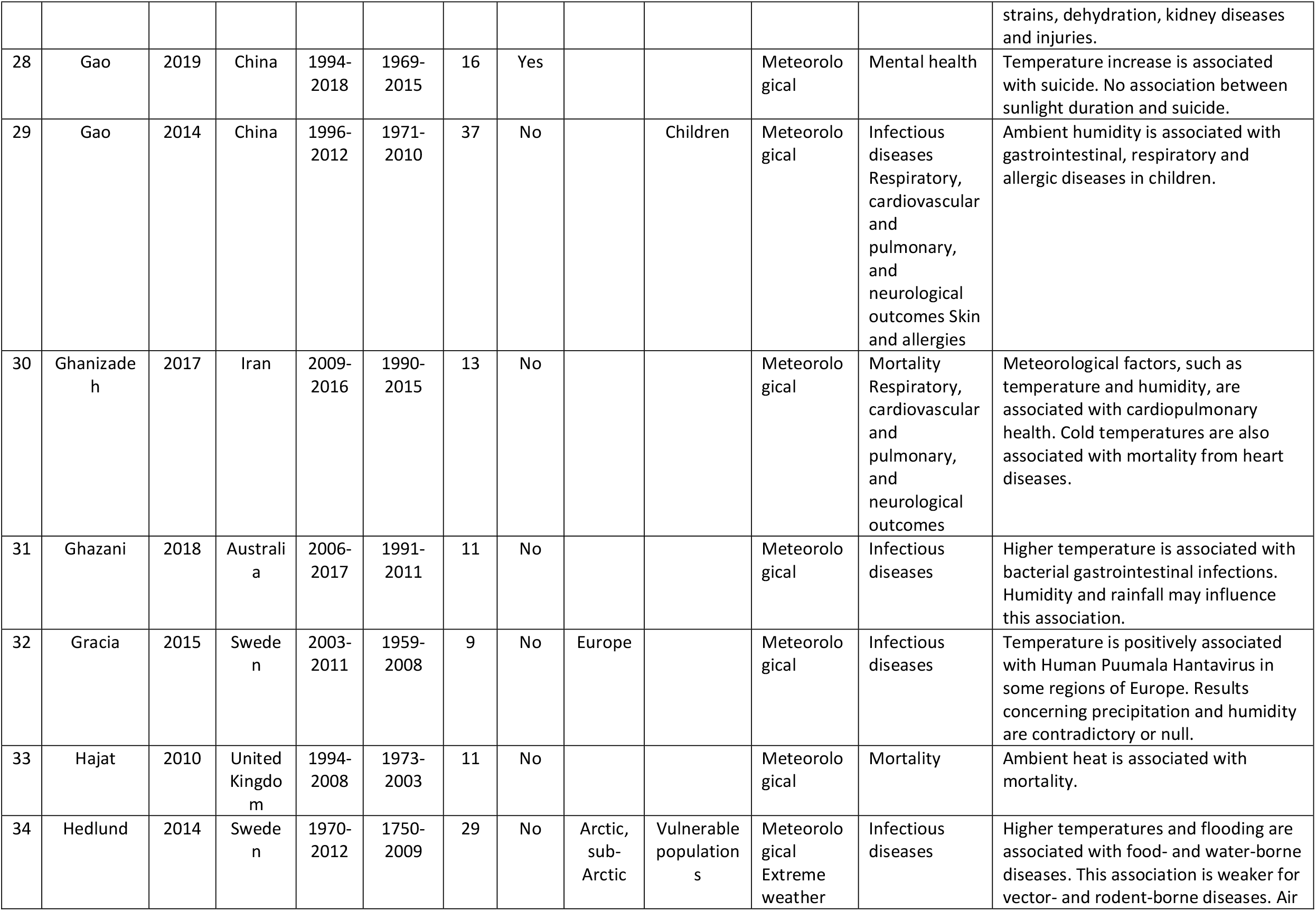
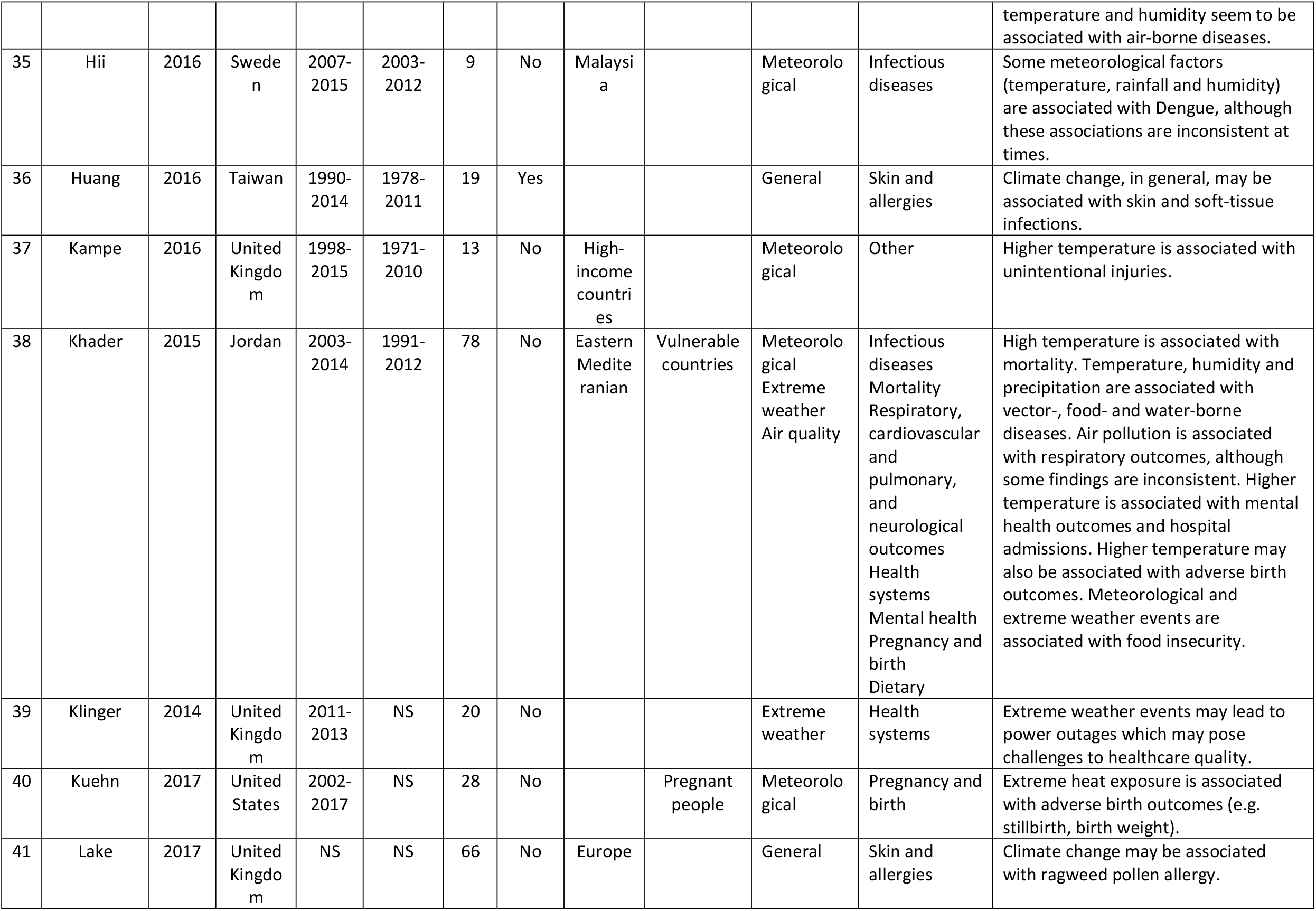
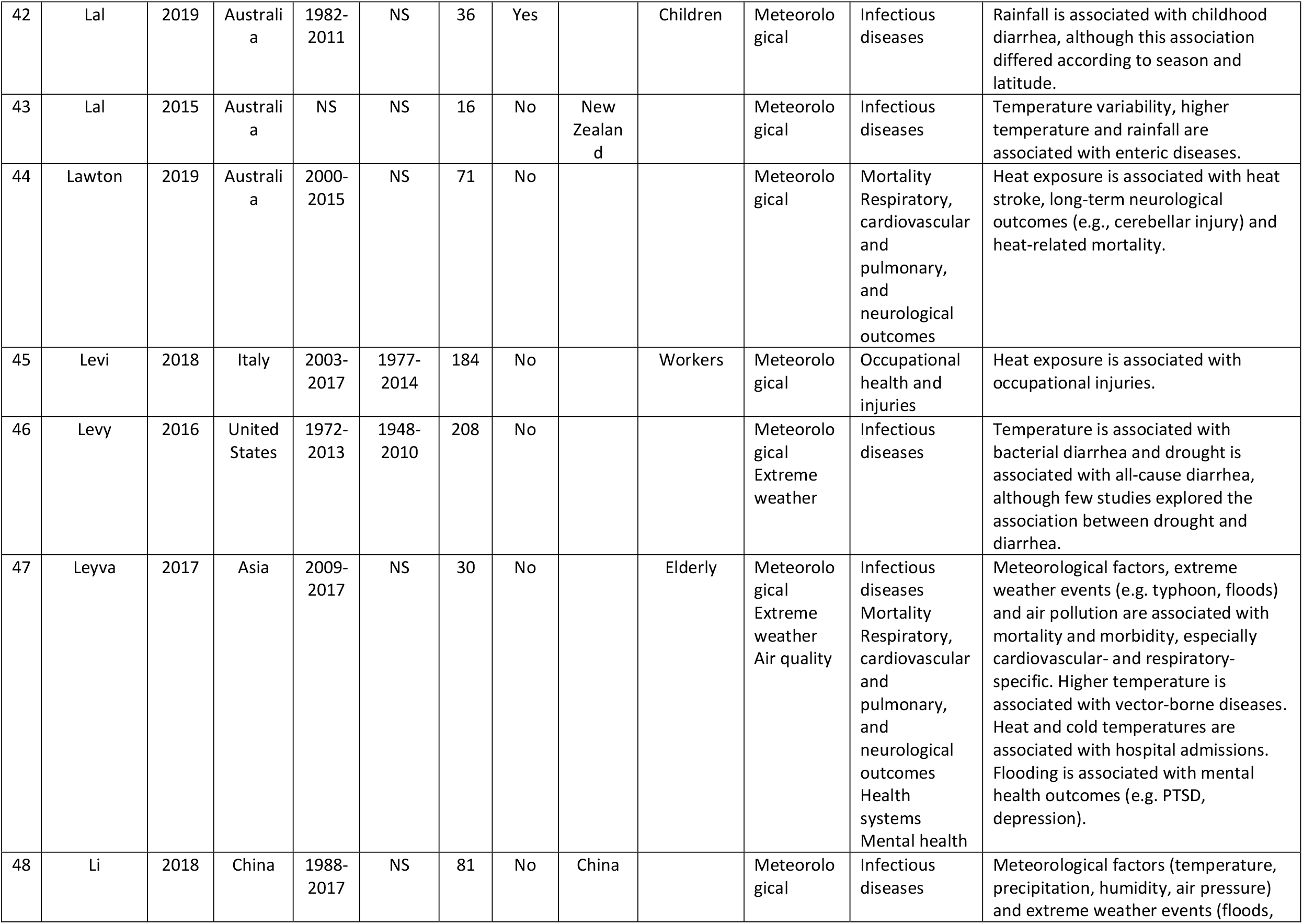
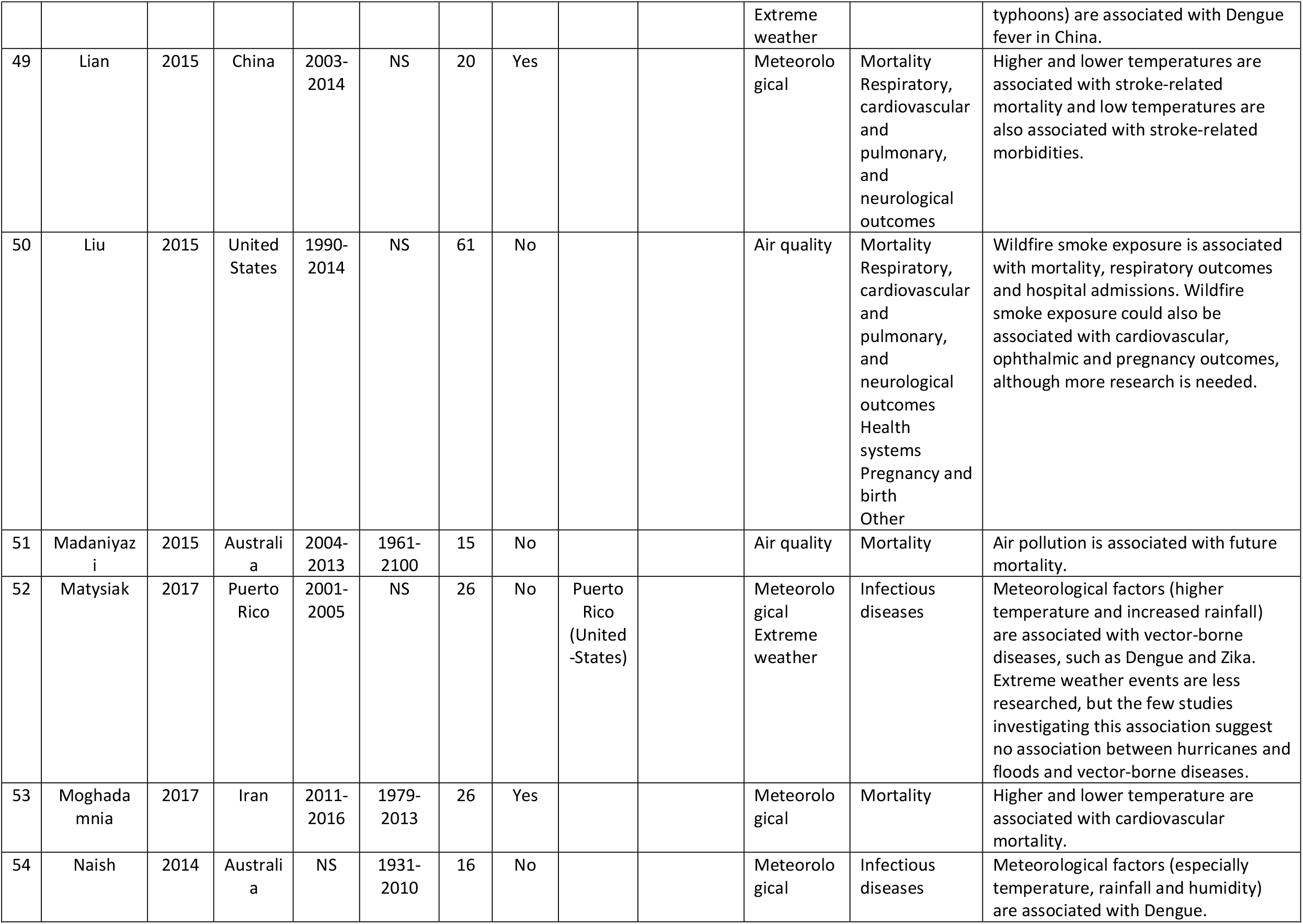
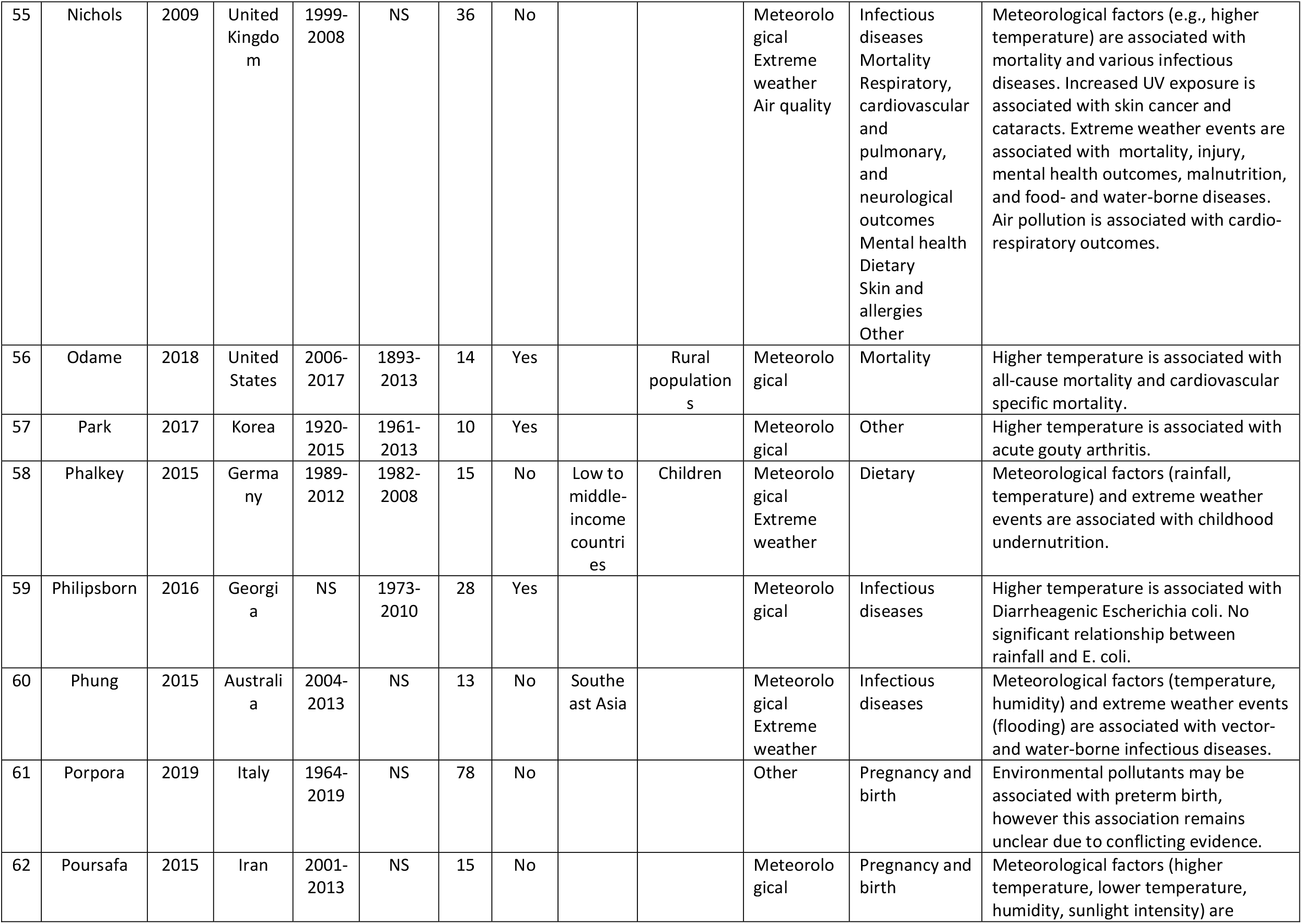
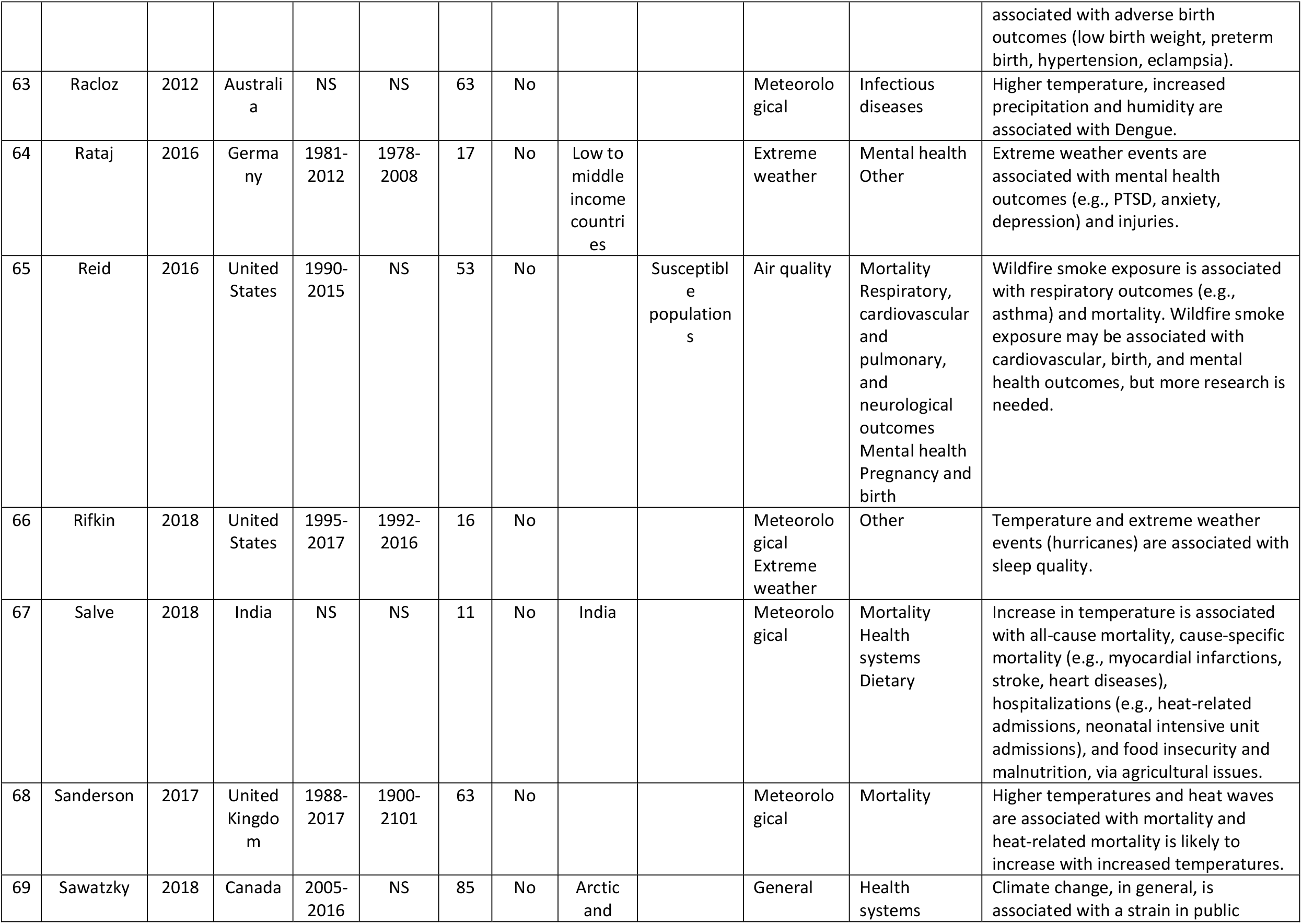
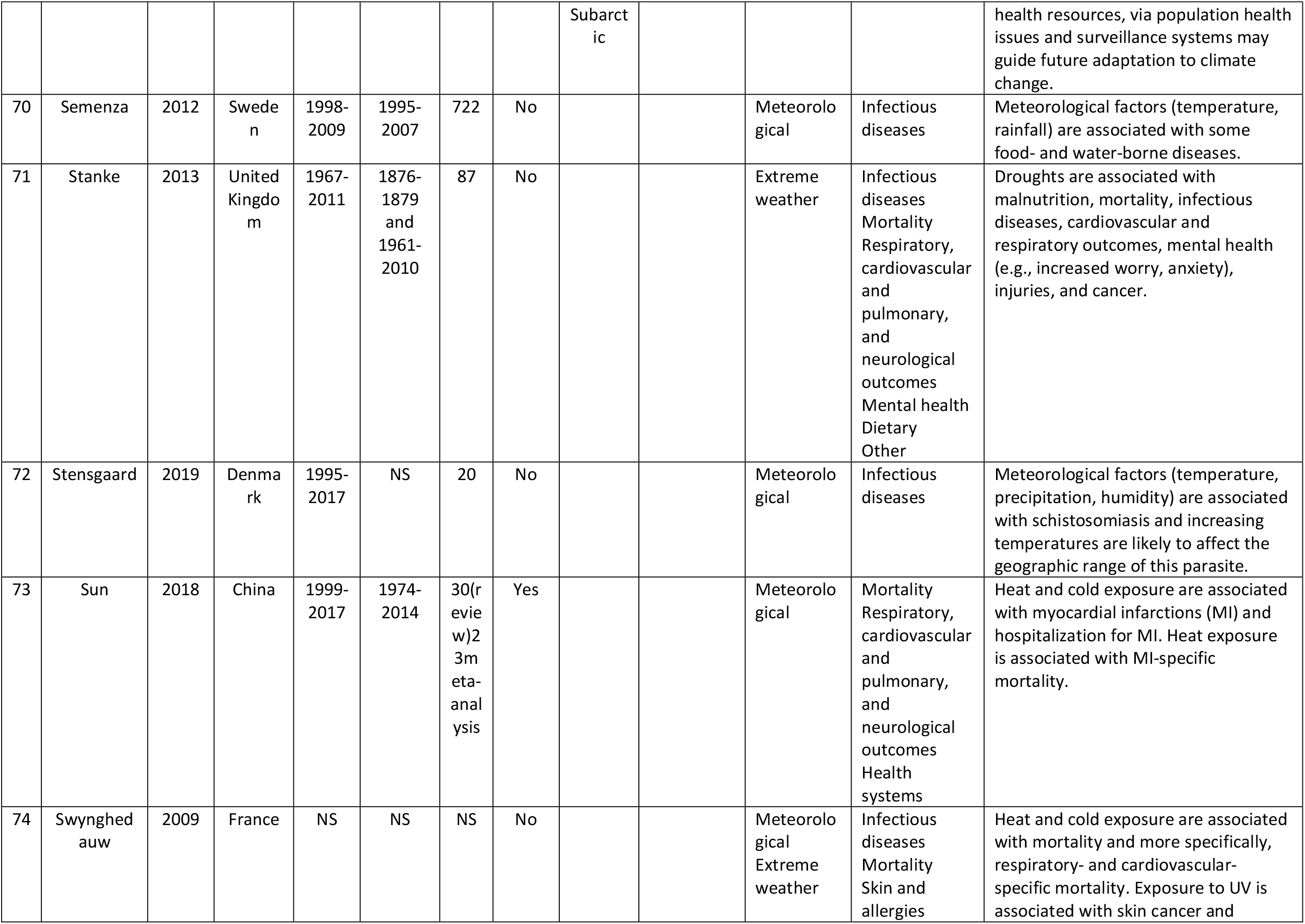
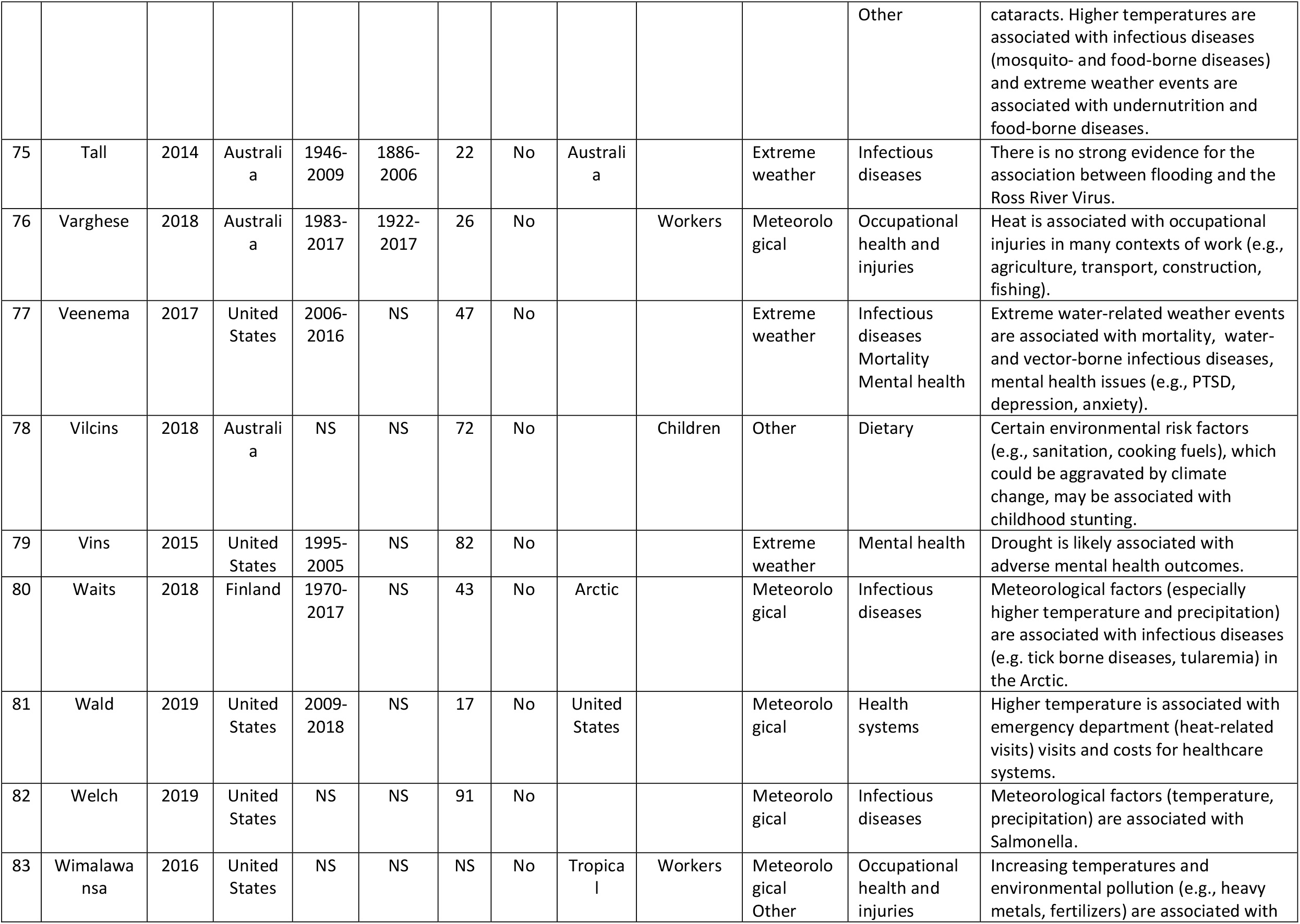
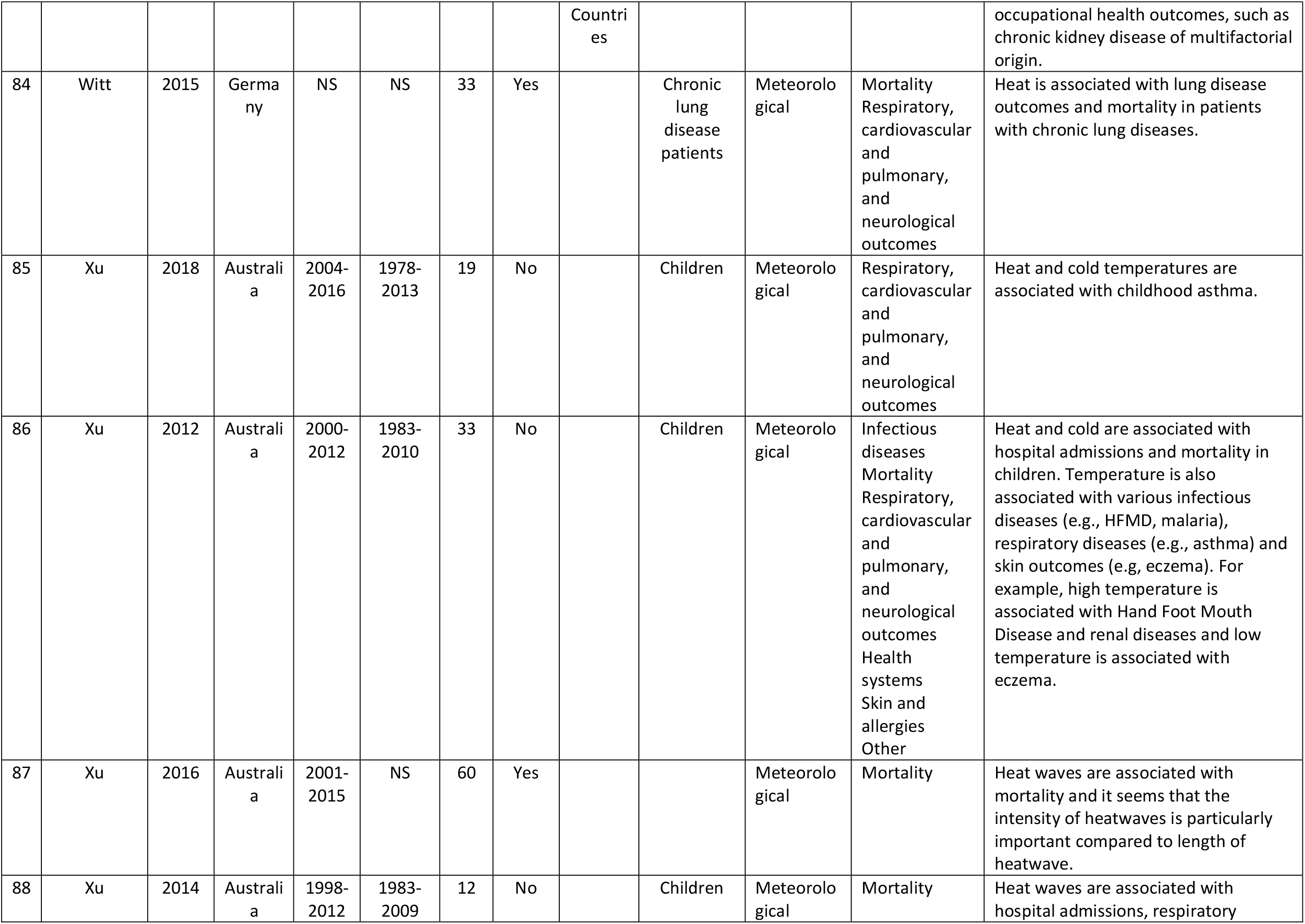
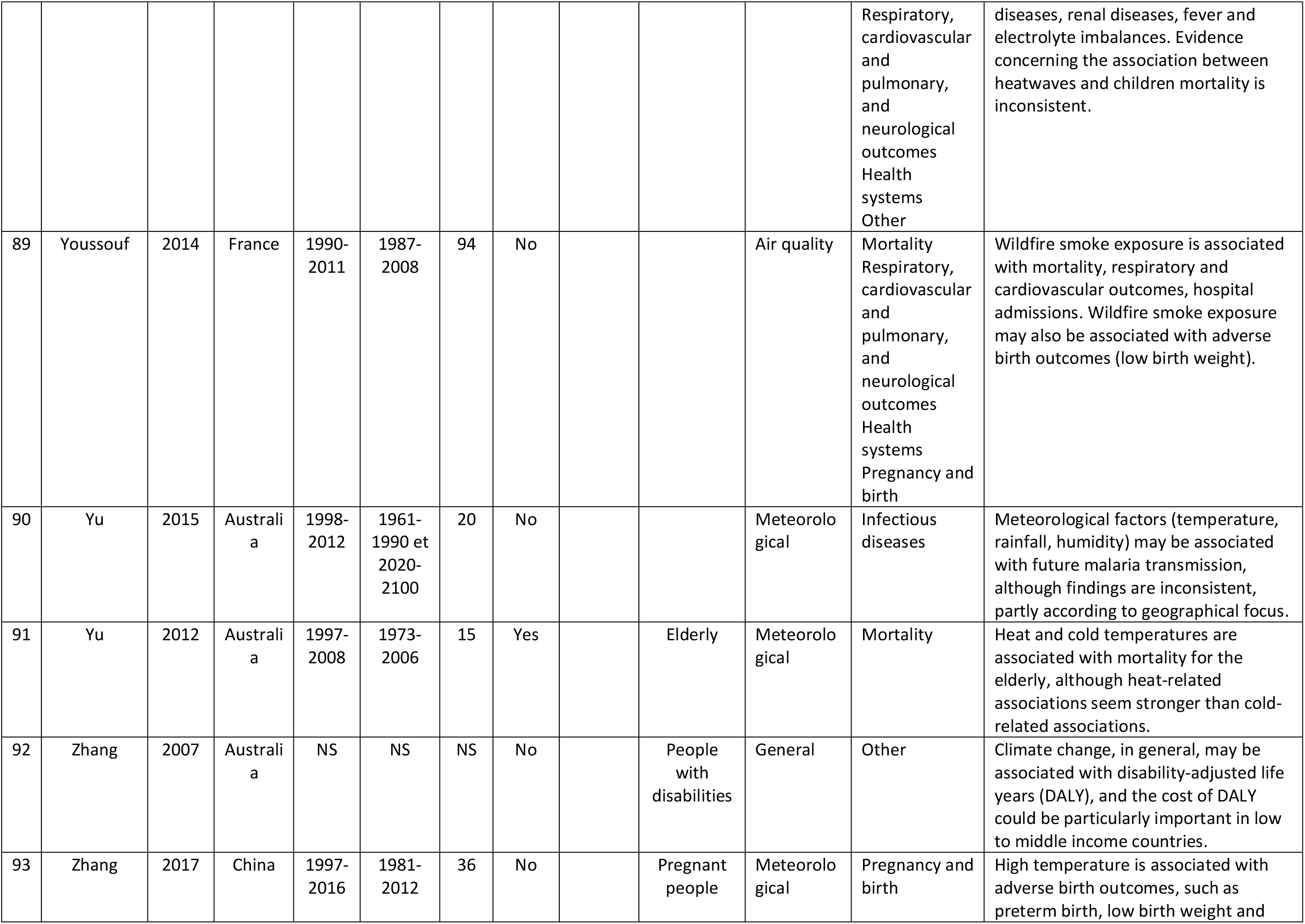
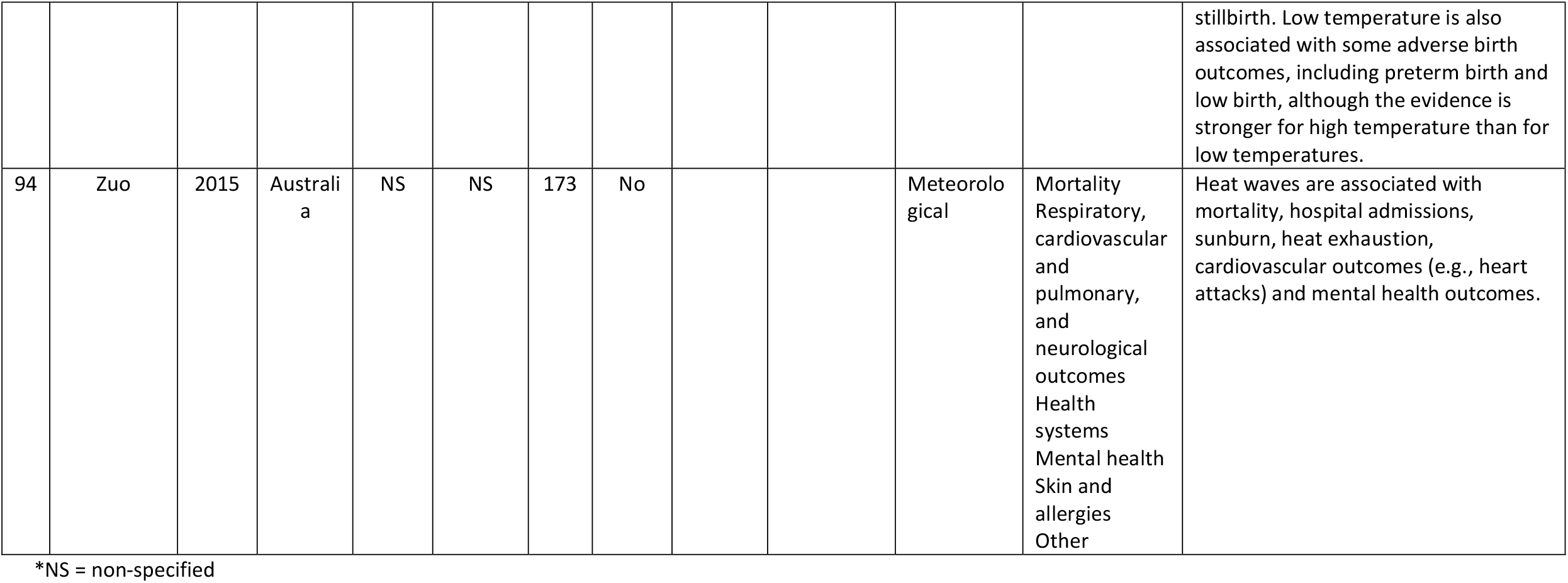

|  | First Author | Year | Country | Years of included publications | Years of the studies included in the reviews | # of articles | Meta-analysis | Specific Area of focus | Specific Population of interest | Climate Impact | Health Outcome | Summary of findings |
| --- | --- | --- | --- | --- | --- | --- | --- | --- | --- | --- | --- | --- |
| 1 | Alderman | 2012 | Australia | 2004-2011 | 1931-2007 | 35 | No |  |  | Extreme weather | Infectious diseases<br>Mortality<br>Health systems<br>Mental health<br>Pregnancy and birth<br>Other | Floods are associated with infectious diseases (water- and vector-borne diseases), mortality (e.g., drowning), exacerbation of chronic illnesses, mental health issues (e.g., PTSD), hospital admissions, pregnancy outcomes and injuries. |
| 2 | Amegah | 2016 | Ghana | 1995-2014 | 1960-2010 | 23 | No | Sub-saharan Africa |  | Meteorological | Infectious diseases<br>Mortality<br>Respiratory, cardiovascular and pulmonary, and neurological outcomes<br>Health systems<br>Dietary<br>Skin and allergies | Temperature and temperature variability are suggested to be associated with infectious diseases, such as Ebola, cardiovascular diseases, mortality, cholera outbreaks, meningitis, undernutrition in children, respiratory outcomes, such as asthma, and skin diseases. |
| 3 | An | 2018 | United States | 2002-2017 | 2002-2016 | 50 | No |  |  | General | Dietary | Climate change is associated with obesity. This association can be explained in 4 ways, such that climate change may impact obesity, obesity may impact climate change, both factors may be associated with common causes, or both factors may influence each other. |
| 4 | Augustin | 2008 | Germany | 1996-2006 | NS | 320 | No | Germany |  | Meteorological | Skin and allergies | Although skin and allergic diseases are climate sensitive, there is not sufficient evidence to suggest a prediction concerning skin and allergic diseases linked to climate change in Germany. |
| 5 | Babaie | 2018 | Iran | 2007-2017 | 1970-2015 | 14 | No | Iran |  | Meteorological | Infectious diseases | Temperature, precipitation and humidity are associated with the risk of transmission of Malaria. |
| 6 | Bai | 2013 | China | 1995-2011 | 1951-2010 | 57 | No | China |  | Meteorological | Infectious diseases | Variability in temperature, precipitation and wind are associated with the risk of transmission of mosquito-borne diseases. |
| 7 | Benevolenza | 2019 | United States | 2006-2017 | 2005-2015 | 13 | No |  | Vulnerable populations | Extreme weather | Mental health<br>Pregnancy and birth<br>Other | Extreme weather events are associated with an exacerbation of pre-existing chronic health conditions, mental health issues (e.g., PTSD, isolation) and adverse birth outcomes. |
| 8 | Berhane | 2016 | Ethiopia | NS | NS | 23 | No | Ethiopia |  | Meteorological<br>Extreme weather | Infectious diseases<br>Dietary | Meteorological factors and extreme weather events are associated with under- and mal- nutrition and the increased risk of climate sensitive infectious diseases (e.g., malaria, diarrhea, zoonotic infections, etc.). |
| 9 | Bernhardt | 2019 | Germany | 1997-2017 | NS | 464 | No |  |  | Meteorological | Infectious diseases | Rising temperatures are predicted to be associated with myiasis in the future. |
| 10 | Binazzi | 2019 | Italy | NS | 1994-2013 | 8 | Yes |  | Workers | Meteorological | Occupational health and injuries | High temperatures are positively associated with occupational injuries. |
| 11 | Bonafede | 2016 | Italy | 2000-2014 | 1985-2010 | 8 | No |  | Workers | Meteorological | Occupational health and injuries | Extreme temperature (particularly heat) is associated with occupational injuries. |
| 12 | Brown | 2013 | United Kingdom | 2004-2012 | 1975-2012 | 38 | No | Europe |  | Extreme weather | Infectious diseases | Floods are associated with infectious diseases, including water-, rodent- and vector-borne diseases (from weeks to months after flooding). |
| 13 | Bunker | 2016 | Germany | 1995-2015 | 1974-2013 | 61 | Yes |  | Elderly | Meteorological | Mortality<br>Respiratory, cardiovascular and pulmonary, and neurological outcomes<br>Other | Higher and lower temperatures are associated with cardiovascular and respiratory morbidity and mortality. Heat is also associated with diabetes and genitourinary infections. |
| 14 | Campbell | 2018 | Australia | 1964-2017 | NS | 188 | No |  |  | Meteorological | Mortality<br>Health systems | Most studies exploring the heat impacts on health focus on mortality outcomes, followed by hospital admissions and ambulance call outs. |
| 15 | Cann | 2013 | United Kingdom | 1973-2010 | NS | 83 | No |  |  | Extreme weather | Infectious diseases | Extreme water-related events are associated with outbreaks of water-related infectious diseases. |
| 16 | Carolan-Olah | 2014 | Australia | 1997-2012 | 1988-2009 | 7 | No |  |  | Meteorological | Pregnancy and birth | High temperatures are associated with preterm birth. |
| 17 | Cheng | 2014 | China | 1990-2013 | 1941-2012 | 25 | No |  | Adults, Elderly, Children | Meteorological | Infectious diseases<br>Mortality<br>Respiratory, cardiovascular and pulmonary, and neurological outcomes<br>Health systems<br>Other | Diurnal temperature range is associated with mortality and cardiovascular and respiratory outcomes, hospital admissions, and Hand Foot Mouth disease in children and genitourinary outcomes among the elderly. |
| 18 | Coates | 2019 | Denmark | 2003-2018 | NS | 72 | No |  |  | Meteorological | Infectious diseases | Some meteorological factors (temperature, humidity) are positively associated with Hand Foot Mouth disease (HFMD). Precipitation, wind speed and sunshine are not associated with HFMD. |
| 19 | Cong | 2017 | China | 1994-2015 | 1982-2013 | 26 | Yes |  |  | Meteorological | Respiratory, cardiovascular and pulmonary, and neurological outcomes | Temperature drop is associated with asthma. |
| 20 | Cunrui | 2011 | Australia | 1997-2010 | 1961-2100 | 14 | No |  |  | Meteorological | Mortality | Higher temperature is associated with heat-related mortality. |
| 21 | deSousa | 2018 | Brazil | 1976-2016 | NS | 106 | No |  |  | Meteorological | Infectious diseases<br>Respiratory, cardiovascular and pulmonary, and neurological outcomes | Meteorological factors (e.g., temperature, precipitation) are associated with infectious diseases (e.g., dengue, malaria) and cardiovascular and respiratory outcomes. |
| 22 | Dhimal | 2015 | Nepal | 1956-2014 | 1948-2098 | 50 | No | Nepal |  | Meteorological | Infectious diseases | Higher temperatures are associated with the distribution of vector-borne diseases. |
| 23 | Doocy | 2013 | United States | 1975-2011 | 1974-2008 | 60 | No |  |  | Extreme weather | Mortality<br>Other | Cyclones are associated with mortality (e.g., drowning) and injuries. |
| 24 | Duan | 2019 | China | 2010-2018 | 2000-2016 | 51 | Yes | Southeast and East Asia |  | Meteorological | Infectious diseases | Some meteorological factors (mean maximum temperature, rainfall, humidity and sunshine) are positively associated with Hand Foot Mouth disease (HFMD), whereas air pressure is negatively associated with this disease and wind speed is not associated with HFMD. |
| 25 | Fan | 2015 | China | 2004-2013 | 1985-2012 | 33 | Yes |  |  | Meteorological | Infectious diseases | Temperature is positively associated with transmission and incidence of Dengue. |
| 26 | Fernandez | 2015 | Australia | 1995-2014 | NS | 83 | No |  |  | Extreme weather | Mental health | Floods are associated with negative mental health outcomes (e.g., PTSD, increased anxiety, depression, use of psychotropic medication). Conflicting evidence concerning suicide, tobacco, alcohol and substance abuse. |
| 27 | Flouris | 2018 | Greece | 1954-2018 | NS | 111 | Yes |  | Workers | Meteorological | Occupational health and injuries | High temperatures are positively associated with occupational heat strains, dehydration, kidney diseases and injuries. |
| 28 | Gao | 2019 | China | 1994-2018 | 1969-2015 | 16 | Yes |  |  | Meteorological | Mental health | Temperature increase is associated with suicide. No association between sunlight duration and suicide. |
| 29 | Gao | 2014 | China | 1996-2012 | 1971-2010 | 37 | No |  | Children | Meteorological | Infectious diseases<br>Respiratory, cardiovascular and pulmonary, and neurological outcomes<br>Skin and allergies | Ambient humidity is associated with gastrointestinal, respiratory and allergic diseases in children. |
| 30 | Ghanizadeh | 2017 | Iran | 2009-2016 | 1990-2015 | 13 | No |  |  | Meteorological | Mortality<br>Respiratory, cardiovascular and pulmonary, and neurological outcomes | Meteorological factors, such as temperature and humidity, are associated with cardiopulmonary health. Cold temperatures are also associated with mortality from heart diseases. |
| 31 | Ghazani | 2018 | Australia | 2006-2017 | 1991-2011 | 11 | No |  |  | Meteorological | Infectious diseases | Higher temperature is associated with bacterial gastrointestinal infections. Humidity and rainfall may influence this association. |
| 32 | Gracia | 2015 | Sweden | 2003-2011 | 1959-2008 | 9 | No | Europe |  | Meteorological | Infectious diseases | Temperature is positively associated with Human Puumala Hantavirus in some regions of Europe. Results concerning precipitation and humidity are contradictory or null. |
| 33 | Hajat | 2010 | United Kingdom | 1994-2008 | 1973-2003 | 11 | No |  |  | Meteorological | Mortality | Ambient heat is associated with mortality. |
| 34 | Hedlund | 2014 | Sweden | 1970-2012 | 1750-2009 | 29 | No | Arctic, sub-Arctic | Vulnerable populations | Meteorological<br>Extreme weather | Infectious diseases | Higher temperatures and flooding are associated with food- and water-borne diseases. This association is weaker for vector- and rodent-borne diseases. Air temperature and humidity seem to be associated with air-borne diseases. |
| 35 | Hii | 2016 | Sweden | 2007-2015 | 2003-2012 | 9 | No | Malaysia |  | Meteorological | Infectious diseases | Some meteorological factors (temperature, rainfall and humidity) are associated with Dengue, although these associations are inconsistent at times. |
| 36 | Huang | 2016 | Taiwan | 1990-2014 | 1978-2011 | 19 | Yes |  |  | General | Skin and allergies | Climate change, in general, may be associated with skin and soft-tissue infections. |
| 37 | Kampe | 2016 | United Kingdom | 1998-2015 | 1971-2010 | 13 | No | High-income countries |  | Meteorological | Other | Higher temperature is associated with unintentional injuries. |
| 38 | Khader | 2015 | Jordan | 2003-2014 | 1991-2012 | 78 | No | Eastern Mediterranean | Vulnerable countries | Meteorological<br>Extreme weather<br>Air quality | Infectious diseases<br>Mortality<br>Respiratory, cardiovascular and pulmonary, and neurological outcomes<br>Health systems<br>Mental health<br>Pregnancy and birth<br>Dietary | High temperature is associated with mortality. Temperature, humidity and precipitation are associated with vector-, food- and water-borne diseases. Air pollution is associated with respiratory outcomes, although some findings are inconsistent. Higher temperature is associated with mental health outcomes and hospital admissions. Higher temperature may also be associated with adverse birth outcomes. Meteorological and extreme weather events are associated with food insecurity. |
| 39 | Klinger | 2014 | United Kingdom | 2011-2013 | NS | 20 | No |  |  | Extreme weather | Health systems | Extreme weather events may lead to power outages which may pose challenges to healthcare quality. |
| 40 | Kuehn | 2017 | United States | 2002-2017 | NS | 28 | No |  | Pregnant people | Meteorological | Pregnancy and birth | Extreme heat exposure is associated with adverse birth outcomes (e.g. stillbirth, birth weight). |
| 41 | Lake | 2017 | United Kingdom | NS | NS | 66 | No | Europe |  | General | Skin and allergies | Climate change may be associated with ragweed pollen allergy. |
| 42 | Lal | 2019 | Australia | 1982-2011 | NS | 36 | Yes |  | Children | Meteorological | Infectious diseases | Rainfall is associated with childhood diarrhea, although this association differed according to season and latitude. |
| 43 | Lal | 2015 | Australia | NS | NS | 16 | No | New Zealand |  | Meteorological | Infectious diseases | Temperature variability, higher temperature and rainfall are associated with enteric diseases. |
| 44 | Lawton | 2019 | Australia | 2000-2015 | NS | 71 | No |  |  | Meteorological | Mortality<br>Respiratory, cardiovascular and pulmonary, and neurological outcomes | Heat exposure is associated with heat stroke, long-term neurological outcomes (e.g., cerebellar injury) and heat-related mortality. |
| 45 | Levi | 2018 | Italy | 2003-2017 | 1977-2014 | 184 | No |  | Workers | Meteorological | Occupational health and injuries | Heat exposure is associated with occupational injuries. |
| 46 | Levy | 2016 | United States | 1972-2013 | 1948-2010 | 208 | No |  |  | Meteorological<br>Extreme weather | Infectious diseases | Temperature is associated with bacterial diarrhea and drought is associated with all-cause diarrhea, although few studies explored the association between drought and diarrhea. |
| 47 | Leyva | 2017 | Asia | 2009-2017 | NS | 30 | No |  | Elderly | Meteorological<br>Extreme weather<br>Air quality | Infectious diseases<br>Mortality<br>Respiratory, cardiovascular and pulmonary, and neurological outcomes<br>Health systems<br>Mental health | Meteorological factors, extreme weather events (e.g. typhoon, floods) and air pollution are associated with mortality and morbidity, especially cardiovascular- and respiratory-specific. Higher temperature is associated with vector-borne diseases. Heat and cold temperatures are associated with hospital admissions. Flooding is associated with mental health outcomes (e.g. PTSD, depression). |
| 48 | Li | 2018 | China | 1988-2017 | NS | 81 | No | China |  | Meteorological | Infectious diseases | Meteorological factors (temperature, precipitation, humidity, air pressure) and extreme weather events (floods, |
|  |  |  |  |  |  |  |  |  |  | Extreme weather |  | typhoons) are associated with Dengue fever in China. |
| 49 | Lian | 2015 | China | 2003-2014 | NS | 20 | Yes |  |  | Meteorological | Mortality<br>Respiratory, cardiovascular and pulmonary, and neurological outcomes | Higher and lower temperatures are associated with stroke-related mortality and low temperatures are also associated with stroke-related morbidities. |
| 50 | Liu | 2015 | United States | 1990-2014 | NS | 61 | No |  |  | Air quality | Mortality<br>Respiratory, cardiovascular and pulmonary, and neurological outcomes<br>Health systems<br>Pregnancy and birth<br>Other | Wildfire smoke exposure is associated with mortality, respiratory outcomes and hospital admissions. Wildfire smoke exposure could also be associated with cardiovascular, ophthalmic and pregnancy outcomes, although more research is needed. |
| 51 | Madaniyazi | 2015 | Australia | 2004-2013 | 1961-2100 | 15 | No |  |  | Air quality | Mortality | Air pollution is associated with future mortality. |
| 52 | Matysiak | 2017 | Puerto Rico | 2001-2005 | NS | 26 | No | Puerto Rico (United States) |  | Meteorological<br>Extreme weather | Infectious diseases | Meteorological factors (higher temperature and increased rainfall) are associated with vector-borne diseases, such as Dengue and Zika. Extreme weather events are less researched, but the few studies investigating this association suggest no association between hurricanes and floods and vector-borne diseases. |
| 53 | Moghaddamnia | 2017 | Iran | 2011-2016 | 1979-2013 | 26 | Yes |  |  | Meteorological | Mortality | Higher and lower temperature are associated with cardiovascular mortality. |
| 54 | Naish | 2014 | Australia | NS | 1931-2010 | 16 | No |  |  | Meteorological | Infectious diseases | Meteorological factors (especially temperature, rainfall and humidity) are associated with Dengue. |
| 55 | Nichols | 2009 | United Kingdom | 1999-2008 | NS | 36 | No |  |  | Meteorological<br>Extreme weather<br>Air quality | Infectious diseases<br>Mortality<br>Respiratory, cardiovascular and pulmonary, and neurological outcomes<br>Mental health<br>Dietary<br>Skin and allergies<br>Other | Meteorological factors (e.g., higher temperature) are associated with mortality and various infectious diseases. Increased UV exposure is associated with skin cancer and cataracts. Extreme weather events are associated with mortality, injury, mental health outcomes, malnutrition, and food- and water-borne diseases. Air pollution is associated with cardio-respiratory outcomes. |
| 56 | Odame | 2018 | United States | 2006-2017 | 1893-2013 | 14 | Yes |  | Rural populations | Meteorological | Mortality | Higher temperature is associated with all-cause mortality and cardiovascular specific mortality. |
| 57 | Park | 2017 | Korea | 1920-2015 | 1961-2013 | 10 | Yes |  |  | Meteorological | Other | Higher temperature is associated with acute gouty arthritis. |
| 58 | Phalkey | 2015 | Germany | 1989-2012 | 1982-2008 | 15 | No | Low to middle-income countries | Children | Meteorological<br>Extreme weather | Dietary | Meteorological factors (rainfall, temperature) and extreme weather events are associated with childhood undernutrition. |
| 59 | Philipsborn | 2016 | Georgia | NS | 1973-2010 | 28 | Yes |  |  | Meteorological | Infectious diseases | Higher temperature is associated with Diarrheagenic Escherichia coli. No significant relationship between rainfall and E. coli. |
| 60 | Phung | 2015 | Australia | 2004-2013 | NS | 13 | No | Southeast Asia |  | Meteorological<br>Extreme weather | Infectious diseases | Meteorological factors (temperature, humidity) and extreme weather events (flooding) are associated with vector- and water-borne infectious diseases. |
| 61 | Porpora | 2019 | Italy | 1964-2019 | NS | 78 | No |  |  | Other | Pregnancy and birth | Environmental pollutants may be associated with preterm birth, however this association remains unclear due to conflicting evidence. |
| 62 | Poursafa | 2015 | Iran | 2001-2013 | NS | 15 | No |  |  | Meteorological | Pregnancy and birth | Meteorological factors (higher temperature, lower temperature, humidity, sunlight intensity) are associated with adverse birth outcomes (low birth weight, preterm birth, hypertension, eclampsia). |
| 63 | Racloz | 2012 | Australia | NS | NS | 63 | No |  |  | Meteorological | Infectious diseases | Higher temperature, increased precipitation and humidity are associated with Dengue. |
| 64 | Rataj | 2016 | Germany | 1981-2012 | 1978-2008 | 17 | No | Low to middle income countries |  | Extreme weather | Mental health<br>Other | Extreme weather events are associated with mental health outcomes (e.g., PTSD, anxiety, depression) and injuries. |
| 65 | Reid | 2016 | United States | 1990-2015 | NS | 53 | No |  | Susceptible populations | Air quality | Mortality<br>Respiratory, cardiovascular and pulmonary, and neurological outcomes<br>Mental health<br>Pregnancy and birth | Wildfire smoke exposure is associated with respiratory outcomes (e.g., asthma) and mortality. Wildfire smoke exposure may be associated with cardiovascular, birth, and mental health outcomes, but more research is needed. |
| 66 | Rifkin | 2018 | United States | 1995-2017 | 1992-2016 | 16 | No |  |  | Meteorological<br>Extreme weather | Other | Temperature and extreme weather events (hurricanes) are associated with sleep quality. |
| 67 | Salve | 2018 | India | NS | NS | 11 | No | India |  | Meteorological | Mortality<br>Health systems<br>Dietary | Increase in temperature is associated with all-cause mortality, cause-specific mortality (e.g., myocardial infarctions, stroke, heart diseases), hospitalizations (e.g., heat-related admissions, neonatal intensive unit admissions), and food insecurity and malnutrition, via agricultural issues. |
| 68 | Sanderson | 2017 | United Kingdom | 1988-2017 | 1900-2101 | 63 | No |  |  | Meteorological | Mortality | Higher temperatures and heat waves are associated with mortality and heat-related mortality is likely to increase with increased temperatures. |
| 69 | Sawatzky | 2018 | Canada | 2005-2016 | NS | 85 | No | Arctic and |  | General | Health systems | Climate change, in general, is associated with a strain in public |
|  |  |  |  |  |  |  |  | Subarctic |  |  |  | health resources, via population health issues and surveillance systems may guide future adaptation to climate change. |
| 70 | Semenza | 2012 | Sweden | 1998-2009 | 1995-2007 | 722 | No |  |  | Meteorological | Infectious diseases | Meteorological factors (temperature, rainfall) are associated with some food- and water-borne diseases. |
| 71 | Stanke | 2013 | United Kingdom | 1967-2011 | 1876-1879 and 1961-2010 | 87 | No |  |  | Extreme weather | Infectious diseases<br>Mortality<br>Respiratory, cardiovascular and pulmonary, and neurological outcomes<br>Mental health<br>Dietary<br>Other | Droughts are associated with malnutrition, mortality, infectious diseases, cardiovascular and respiratory outcomes, mental health (e.g., increased worry, anxiety), injuries, and cancer. |
| 72 | Stensgaard | 2019 | Denmark | 1995-2017 | NS | 20 | No |  |  | Meteorological | Infectious diseases | Meteorological factors (temperature, precipitation, humidity) are associated with schistosomiasis and increasing temperatures are likely to affect the geographic range of this parasite. |
| 73 | Sun | 2018 | China | 1999-2017 | 1974-2014 | 30(review)23meta-analysis | Yes |  |  | Meteorological | Mortality<br>Respiratory, cardiovascular and pulmonary, and neurological outcomes<br>Health systems | Heat and cold exposure are associated with myocardial infarctions (MI) and hospitalization for MI. Heat exposure is associated with MI-specific mortality. |
| 74 | Swynghedauw | 2009 | France | NS | NS | NS | No |  |  | Meteorological<br>Extreme weather | Infectious diseases<br>Mortality<br>Skin and allergies | Heat and cold exposure are associated with mortality and more specifically, respiratory- and cardiovascular-specific mortality. Exposure to UV is associated with skin cancer and |
|  |  |  |  |  |  |  |  |  |  |  | Other | cataracts. Higher temperatures are associated with infectious diseases (mosquito- and food-borne diseases) and extreme weather events are associated with undernutrition and food-borne diseases. |
| 75 | Tall | 2014 | Australia | 1946-2009 | 1886-2006 | 22 | No | Australia |  | Extreme weather | Infectious diseases | There is no strong evidence for the association between flooding and the Ross River Virus. |
| 76 | Varghese | 2018 | Australia | 1983-2017 | 1922-2017 | 26 | No |  | Workers | Meteorological | Occupational health and injuries | Heat is associated with occupational injuries in many contexts of work (e.g., agriculture, transport, construction, fishing). |
| 77 | Veenema | 2017 | United States | 2006-2016 | NS | 47 | No |  |  | Extreme weather | Infectious diseases<br>Mortality<br>Mental health | Extreme water-related weather events are associated with mortality, water- and vector-borne infectious diseases, mental health issues (e.g., PTSD, depression, anxiety). |
| 78 | Vilcins | 2018 | Australia | NS | NS | 72 | No |  | Children | Other | Dietary | Certain environmental risk factors (e.g., sanitation, cooking fuels), which could be aggravated by climate change, may be associated with childhood stunting. |
| 79 | Vins | 2015 | United States | 1995-2005 | NS | 82 | No |  |  | Extreme weather | Mental health | Drought is likely associated with adverse mental health outcomes. |
| 80 | Waits | 2018 | Finland | 1970-2017 | NS | 43 | No | Arctic |  | Meteorological | Infectious diseases | Meteorological factors (especially higher temperature and precipitation) are associated with infectious diseases (e.g. tick borne diseases, tularemia) in the Arctic. |
| 81 | Wald | 2019 | United States | 2009-2018 | NS | 17 | No | United States |  | Meteorological | Health systems | Higher temperature is associated with emergency department (heat-related visits) visits and costs for healthcare systems. |
| 82 | Welch | 2019 | United States | NS | NS | 91 | No |  |  | Meteorological | Infectious diseases | Meteorological factors (temperature, precipitation) are associated with Salmonella. |
| 83 | Wimalawansa | 2016 | United States | NS | NS | NS | No | Tropical | Workers | Meteorological<br>Other | Occupational health and injuries | Increasing temperatures and environmental pollution (e.g., heavy metals, fertilizers) are associated with |
|  |  |  |  |  |  |  |  | Countries |  |  |  | occupational health outcomes, such as chronic kidney disease of multifactorial origin. |
| 84 | Witt | 2015 | Germany | NS | NS | 33 | Yes |  | Chronic lung disease patients | Meteorological | Mortality<br>Respiratory, cardiovascular and pulmonary, and neurological outcomes | Heat is associated with lung disease outcomes and mortality in patients with chronic lung diseases. |
| 85 | Xu | 2018 | Australia | 2004-2016 | 1978-2013 | 19 | No |  | Children | Meteorological | Respiratory, cardiovascular and pulmonary, and neurological outcomes | Heat and cold temperatures are associated with childhood asthma. |
| 86 | Xu | 2012 | Australia | 2000-2012 | 1983-2010 | 33 | No |  | Children | Meteorological | Infectious diseases<br>Mortality<br>Respiratory, cardiovascular and pulmonary, and neurological outcomes<br>Health systems<br>Skin and allergies<br>Other | Heat and cold are associated with hospital admissions and mortality in children. Temperature is also associated with various infectious diseases (e.g., HFMD, malaria), respiratory diseases (e.g., asthma) and skin outcomes (e.g., eczema). For example, high temperature is associated with Hand Foot Mouth Disease and renal diseases and low temperature is associated with eczema. |
| 87 | Xu | 2016 | Australia | 2001-2015 | NS | 60 | Yes |  |  | Meteorological | Mortality | Heat waves are associated with mortality and it seems that the intensity of heatwaves is particularly important compared to length of heatwave. |
| 88 | Xu | 2014 | Australia | 1998-2012 | 1983-2009 | 12 | No |  | Children | Meteorological | Mortality | Heat waves are associated with hospital admissions, respiratory |
|  |  |  |  |  |  |  |  |  |  |  | Respiratory, cardiovascular and pulmonary, and neurological outcomes<br>Health systems<br>Other | diseases, renal diseases, fever and electrolyte imbalances. Evidence concerning the association between heatwaves and children mortality is inconsistent. |
| 89 | Yousseuf | 2014 | France | 1990-2011 | 1987-2008 | 94 | No |  |  | Air quality | Mortality<br>Respiratory, cardiovascular and pulmonary, and neurological outcomes<br>Health systems<br>Pregnancy and birth | Wildfire smoke exposure is associated with mortality, respiratory and cardiovascular outcomes, hospital admissions. Wildfire smoke exposure may also be associated with adverse birth outcomes (low birth weight). |
| 90 | Yu | 2015 | Australia | 1998-2012 | 1961-1990 et 2020-2100 | 20 | No |  |  | Meteorological | Infectious diseases | Meteorological factors (temperature, rainfall, humidity) may be associated with future malaria transmission, although findings are inconsistent, partly according to geographical focus. |
| 91 | Yu | 2012 | Australia | 1997-2008 | 1973-2006 | 15 | Yes |  | Elderly | Meteorological | Mortality | Heat and cold temperatures are associated with mortality for the elderly, although heat-related associations seem stronger than cold-related associations. |
| 92 | Zhang | 2007 | Australia | NS | NS | NS | No |  | People with disabilities | General | Other | Climate change, in general, may be associated with disability-adjusted life years (DALY), and the cost of DALY could be particularly important in low to middle income countries. |
| 93 | Zhang | 2017 | China | 1997-2016 | 1981-2012 | 36 | No |  | Pregnant people | Meteorological | Pregnancy and birth | High temperature is associated with adverse birth outcomes, such as preterm birth, low birth weight and stillbirth. Low temperature is also associated with some adverse birth outcomes, including preterm birth and low birth, although the evidence is stronger for high temperature than for low temperatures. |
| 94 | Zuo | 2015 | Australia | NS | NS | 173 | No |  |  | Meteorological | Mortality<br>Respiratory, cardiovascular and pulmonary, and neurological outcomes<br>Health systems<br>Mental health<br>Skin and allergies<br>Other | Heat waves are associated with mortality, hospital admissions, sunburn, heat exhaustion, cardiovascular outcomes (e.g., heart attacks) and mental health outcomes. |

\*NS = non-specified

## Appendix 4. Summary of quality assessment according to revised AMSTAR-2 items. (Y = yes, PY = partial yes, N = no, NA = non-applicable).

|  |  | AMSTAR-2 Items |  |  |  |  |  |  |  |  |  |  |  |  |  |  |  |  |
| --- | --- | --- | --- | --- | --- | --- | --- | --- | --- | --- | --- | --- | --- | --- | --- | --- | --- | --- |
| First author | Year | 1 | 1b | 2 | 3 | 4 | 5 | 6 | 7 | 8 | 9 | 10 | 11 | 12 | 13 | 14 | 15 | 16 |
| Alderman | 2012 | Y | N | N | PY | PY | N | N | PY | PY | N | N | NA | NA | Y | N | NA | N |
| Amegah | 2016 | Y | N | N | Y | PY | N | N | PY | Y | Y | N | NA | NA | Y | Y | NA | Y |
| An | 2018 | N | Y | N | N | PY | Y | N | Y | N | Y | N | NA | NA | Y | N | NA | Y |
| Augustin | 2008 | Y | Y | N | N | PY | N | N | PY | N | PY | N | NA | NA | Y | N | NA | Y |
| Babaie | 2018 | Y | Y | N | N | PY | Y | N | Y | PY | N | N | NA | NA | N | N | NA | N |
| Bai | 2013 | Y | Y | N | PY | PY | N | N | PY | PY | Y | N | NA | NA | Y | N | NA | Y |
| Benevolenza | 2019 | Y | Y | N | PY | N | N | N | Y | PY | PY | N | NA | NA | Y | N | NA | N |
| Berhane | 2016 | Y | N | Y | N | N | N | N | N | PY | PY | N | NA | NA | Y | N | NA | N |
| Bernhardt | 2019 | Y | N | N | N | N | N | N | PY | PY | PY | N | NA | NA | PY | N | NA | Y |
| Binazzi | 2019 | Y | Y | N | Y | PY | Y | N | Y | Y | Y | N | Y | Y | Y | Y | Y | Y |
| Bonafede | 2016 | Y | N | N | PY | PY | Y | N | Y | Y | Y | N | NA | NA | Y | Y | NA | Y |
| Brown | 2013 | Y | Y | N | PY | PY | N | N | Y | Y | PY | N | NA | NA | Y | Y | NA | Y |
| Bunker | 2016 | Y | Y | N | Y | N | Y | PY | Y | Y | PY | N | Y | Y | Y | Y | Y | Y |
| Campbell | 2018 | Y | N | N | N | PY | N | N | Y | PY | N | N | NA | NA | Y | PY | NA | Y |
| Cann | 2013 | Y | Y | Y | PY | PY | N | N | Y | PY | PY | N | NA | NA | Y | Y | NA | Y |
| Carolan-Olah | 2014 | Y | Y | N | N | PY | N | N | Y | PY | Y | N | NA | NA | PY | PY | NA | N |
| Cheng | 2014 | Y | N | N | PY | PY | N | N | Y | Y | Y | N | NA | NA | Y | PY | NA | N |
| Coates | 2019 | N | N | N | N | N | N | N | PY | PY | Y | N | NA | NA | Y | PY | NA | N |
| Cong | 2017 | N | Y | N | N | PY | N | Y | Y | Y | Y | N | Y | N | Y | Y | Y | Y |
| Cunrui | 2011 | Y | N | N | PY | PY | N | N | Y | PY | N | N | NA | NA | Y | N | NA | Y |
| deSousa | 2018 | Y | N | N | PY | PY | N | N | Y | N | N | N | NA | NA | Y | N | NA | Y |
| Dhimal | 2015 | Y | Y | N | PY | PY | N | N | Y | Y | N | N | NA | NA | N | N | NA | N |
| Doocy | 2013 | Y | N | N | N | PY | Y | N | Y | Y | N | N | NA | NA | Y | PY | NA | Y |
| Duan | 2019 | Y | N | N | N | PY | N | N | Y | PY | Y | N | Y | Y | Y | Y | Y | Y |
| Fan | 2015 | N | Y | N | N | PY | Y | Y | PY | PY | Y | N | Y | Y | Y | Y | Y | Y |
| Fernandez | 2015 | Y | Y | N | PY | PY | Y | N | Y | PY | PY | N | NA | NA | Y | N | NA | N |
| Flouris | 2018 | Y | N | Y | PY | PY | Y | Y | Y | Y | Y | Y | Y | Y | Y | Y | Y | Y |
| Gao | 2019 | N | Y | N | PY | PY | Y | N | Y | Y | Y | N | Y | Y | Y | Y | Y | Y |
| Gao | 2014 | Y | N | N | PY | PY | N | PY | Y | Y | Y | N | NA | NA | Y | Y | NA | Y |
| Ghanizadeh | 2017 | N | Y | N | PY | N | Y | PY | Y | PY | Y | N | NA | NA | N | N | NA | Y |
| Ghazani | 2018 | N | N | N | N | PY | Y | N | Y | Y | PY | N | NA | NA | Y | PY | NA | Y |
| Gracia | 2015 | Y | Y | N | PY | PY | Y | N | Y | PY | Y | N | NA | NA | Y | N | NA | Y |
| Hajat | 2010 | Y | N | N | Y | PY | N | N | Y | PY | N | N | NA | NA | N | PY | NA | Y |
| Hedlund | 2014 | Y | N | N | PY | N | Y | Y | Y | PY | Y | N | NA | NA | Y | N | NA | Y |
| Hii | 2016 | Y | N | N | PY | PY | N | N | PY | PY | N | N | NA | NA | N | N | NA | Y |
| Huang | 2016 | N | N | N | N | PY | Y | N | PY | Y | Y | N | Y | Y | Y | PY | N | Y |
| Kampe | 2016 | Y | N | N | PY | PY | Y | N | Y | Y | PY | N | NA | NA | Y | N | NA | Y |
| Khader | 2015 | Y | N | N | PY | PY | Y | N | Y | Y | N | N | NA | NA | Y | PY | NA | Y |
| Klinger | 2014 | N | N | N | N | PY | Y | N | Y | N | Y | N | NA | NA | Y | N | NA | Y |
| Kuehn | 2017 | Y | N | N | N | PY | N | N | Y | Y | PY | N | NA | NA | Y | N | NA | Y |
| Lake | 2017 | Y | Y | N | N | PY | Y | Y | PY | PY | N | N | NA | NA | N | N | NA | Y |
| Lal | 2019 | Y | N | Y | N | PY | N | Y | Y | PY | Y | N | Y | Y | Y | Y | N | Y |
| Lal | 2015 | Y | Y | N | N | PY | N | N | Y | PY | PY | N | NA | NA | Y | PY | NA | Y |
| Lawton | 2019 | Y | N | N | PY | PY | N | N | Y | Y | N | N | NA | NA | Y | N | NA | Y |
| Levi | 2018 | Y | Y | N | PY | PY | Y | N | Y | Y | N | N | NA | NA | Y | PY | NA | Y |
| Levy | 2016 | N | N | N | PY | PY | Y | PY | Y | Y | Y | N | NA | NA | Y | Y | NA | Y |
| Leyva | 2017 | Y | Y | N | PY | PY | N | N | Y | N | Y | N | NA | NA | Y | N | NA | N |
| Li | 2018 | Y | N | N | N | PY | N | N | N | PY | N | N | NA | NA | N | N | NA | Y |
| Lian | 2015 | Y | N | N | PY | PY | Y | Y | Y | N | Y | N | Y | Y | Y | Y | Y | Y |
| Liu | 2015 | Y | Y | N | Y | PY | N | N | Y | Y | PY | N | NA | NA | Y | PY | NA | Y |
| Madaniyazi | 2015 | N | N | N | PY | PY | N | N | Y | PY | N | N | NA | NA | Y | N | NA | Y |
| Matysiak | 2017 | Y | Y | N | PY | PY | N | N | Y | PY | Y | N | NA | NA | Y | N | NA | Y |
| Moghadamnia | 2017 | Y | N | N | PY | PY | Y | N | Y | Y | Y | N | Y | N | Y | Y | N | Y |
| Naish | 2014 | Y | Y | N | PY | PY | N | N | PY | PY | PY | N | NA | NA | Y | N | NA | Y |
| Nichols | 2009 | Y | N | N | PY | PY | Y | Y | Y | Y | N | N | NA | NA | N | N | NA | Y |
| Odame | 2018 | Y | N | N | PY | PY | N | N | PY | PY | N | N | Y | N | Y | Y | Y | Y |
| Park | 2017 | Y | N | N | N | PY | Y | Y | N | PY | Y | N | Y | PY | Y | Y | N | Y |
| Phalkey | 2015 | Y | Y | N | PY | PY | Y | Y | Y | PY | PY | N | NA | NA | Y | N | NA | Y |
| Philipsborn | 2016 | Y | Y | N | PY | N | N | N | Y | PY | N | N | Y | Y | N | Y | N | Y |
| Phung | 2015 | Y | Y | N | N | PY | N | N | Y | Y | PY | Y | NA | NA | N | N | NA | Y |
| Porpora | 2019 | Y | Y | Y | PY | PY | N | N | Y | PY | N | N | NA | NA | N | Y | NA | Y |
| Poursafa | 2015 | Y | N | N | PY | PY | Y | Y | PY | Y | PY | N | NA | NA | Y | Y | NA | Y |
| Racloz | 2012 | Y | N | N | N | PY | N | N | N | PY | N | N | NA | NA | Y | Y | NA | Y |
| Rataj | 2016 | Y | Y | Y | PY | PY | Y | Y | PY | PY | Y | N | NA | NA | Y | Y | NA | Y |
| Reid | 2016 | Y | N | N | PY | PY | N | N | N | PY | PY | N | NA | NA | Y | PY | NA | Y |
| Rifkin | 2018 | Y | Y | Y | PY | PY | Y | Y | Y | PY | N | N | NA | NA | Y | N | NA | Y |
| Salve | 2018 | Y | Y | N | N | PY | Y | Y | PY | PY | N | N | NA | NA | Y | N | NA | Y |
| Sanderson | 2017 | Y | Y | N | N | PY | Y | N | Y | PY | PY | N | NA | NA | Y | N | NA | Y |
| Sawatzky | 2018 | Y | Y | N | N | PY | Y | N | Y | N | N | N | NA | NA | N | N | NA | Y |
| Semenza | 2012 | N | N | N | N | N | N | N | N | N | Y | N | NA | NA | N | N | NA | N |
| Stanke | 2013 | Y | N | N | PY | PY | N | N | Y | PY | N | N | NA | NA | Y | N | NA | Y |
| Stensgaard | 2019 | Y | Y | N | N | PY | N | Y | N | PY | N | N | NA | NA | N | N | NA | Y |
| Sun | 2018 | Y | Y | N | PY | PY | N | Y | Y | Y | Y | N | Y | Y | Y | Y | Y | Y |
| Swynghedauw | 2009 | Y | Y | N | N | N | N | N | N | N | N | N | NA | NA | N | N | NA | Y |
| Tall | 2014 | Y | N | N | N | PY | N | N | Y | Y | PY | N | NA | NA | Y | N | NA | Y |
| Varghese | 2018 | Y | N | N | N | PY | N | N | Y | Y | N | N | NA | NA | Y | N | NA | Y |
| Veenema | 2017 | Y | N | Y | N | PY | N | N | Y | N | PY | N | NA | NA | N | N | NA | N |
| Vilcins | 2018 | Y | N | Y | N | PY | N | N | Y | N | Y | N | NA | NA | Y | N | NA | Y |
| Vins | 2015 | Y | Y | N | N | PY | N | N | Y | N | N | N | NA | NA | Y | PY | NA | Y |
| Waits | 2018 | Y | Y | N | N | PY | N | N | Y | N | N | N | NA | NA | N | N | NA | Y |
| Wald | 2019 | Y | N | N | N | PY | N | N | Y | Y | N | N | NA | NA | Y | N | NA | N |
| Welch | 2019 | Y | Y | N | N | N | N | N | PY | PY | N | N | NA | NA | N | N | NA | N |
| Wimalawansa | 2016 | Y | N | N | N | N | N | N | N | N | N | N | NA | NA | N | N | NA | Y |
| Witt | 2015 | Y | N | N | N | PY | N | N | Y | N | N | N | N | Y | Y | N | N | Y |
| Xu | 2018 | Y | N | N | PY | PY | N | N | PY | PY | N | N | NA | NA | Y | N | NA | Y |
| Xu | 2012 | Y | N | N | PY | PY | N | N | PY | Y | PY | N | NA | NA | N | N | NA | Y |
| Xu | 2016 | Y | N | N | PY | PY | N | N | Y | Y | N | N | Y | N | Y | Y | N | Y |
| Xu | 2014 | Y | Y | N | N | PY | N | N | Y | Y | N | N | NA | NA | N | Y | NA | N |
| Yousseuf | 2014 | Y | Y | N | PY | PY | N | N | Y | Y | N | N | NA | NA | Y | N | NA | Y |
| Yu | 2015 | N | N | N | PY | PY | N | N | Y | Y | N | N | NA | NA | Y | PY | NA | Y |
| Yu | 2012 | Y | Y | N | PY | N | N | Y | Y | Y | N | N | Y | N | Y | Y | Y | Y |
| Zhang | 2007 | Y | N | N | N | N | N | N | N | N | N | N | NA | NA | Y | N | NA | N |
| Zhang | 2017 | Y | Y | N | PY | PY | N | N | PY | Y | Y | N | NA | NA | Y | Y | NA | Y |
| Zuo | 2015 | Y | Y | N | N | PY | N | N | N | N | N | N | NA | NA | N | N | NA | Y |
| TOTAL | Yes | 81 | 44 | 9 | 5 | 0 | 32 | 17 | 64 | 36 | 31 | 2 | 17 | 12 | 70 | 28 | 11 | 78 |
|  | Partial<br>Yes | 0 | 0 | 0 | 47 | 80 | 0 | 4 | 19 | 41 | 22 | 0 | 0 | 1 | 2 | 15 | 0 | 0 |
|  | No | 13 | 50 | 85 | 42 | 14 | 62 | 73 | 11 | 17 | 41 | 92 | 1 | 5 | 22 | 51 | 7 | 16 |
|  | Not-Applic<br>able | 0 | 0 | 0 | 0 | 0 | 0 | 0 | 0 | 0 | 0 | 0 | 76 | 76 | 0 | 0 | 76 | 0 |

## Appendix 5. Summary table of publications according to climate impact and health outcome according to frequencies and references. with a * explore only this specific combination of climate impact and health outcome and therefore appear only once in this table. Number of studies and references (according to alphabetical order) for each combination of categories are presented in the cells. Empty cells in this Table indicate an absence of studies at this combination of categories.

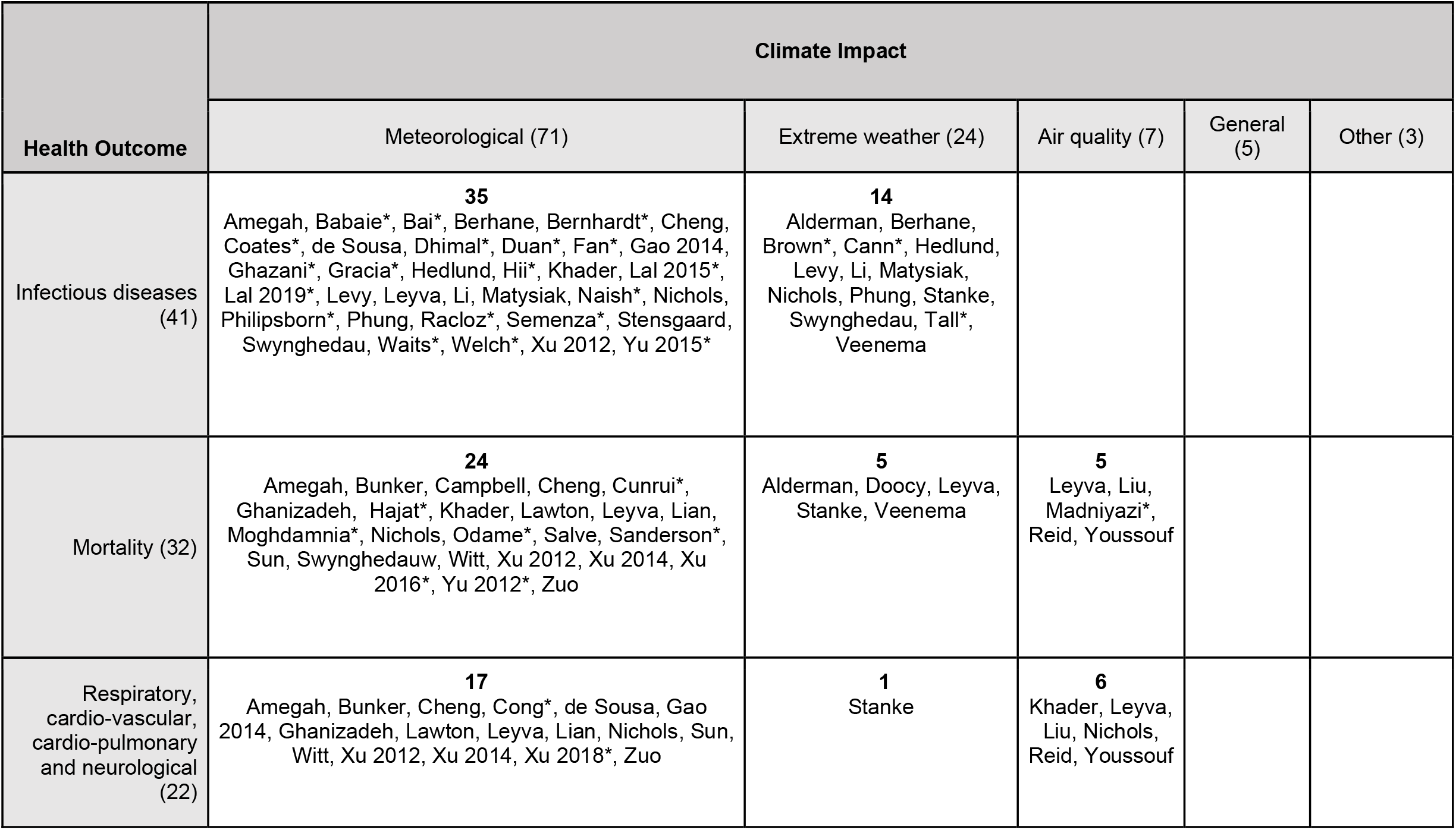
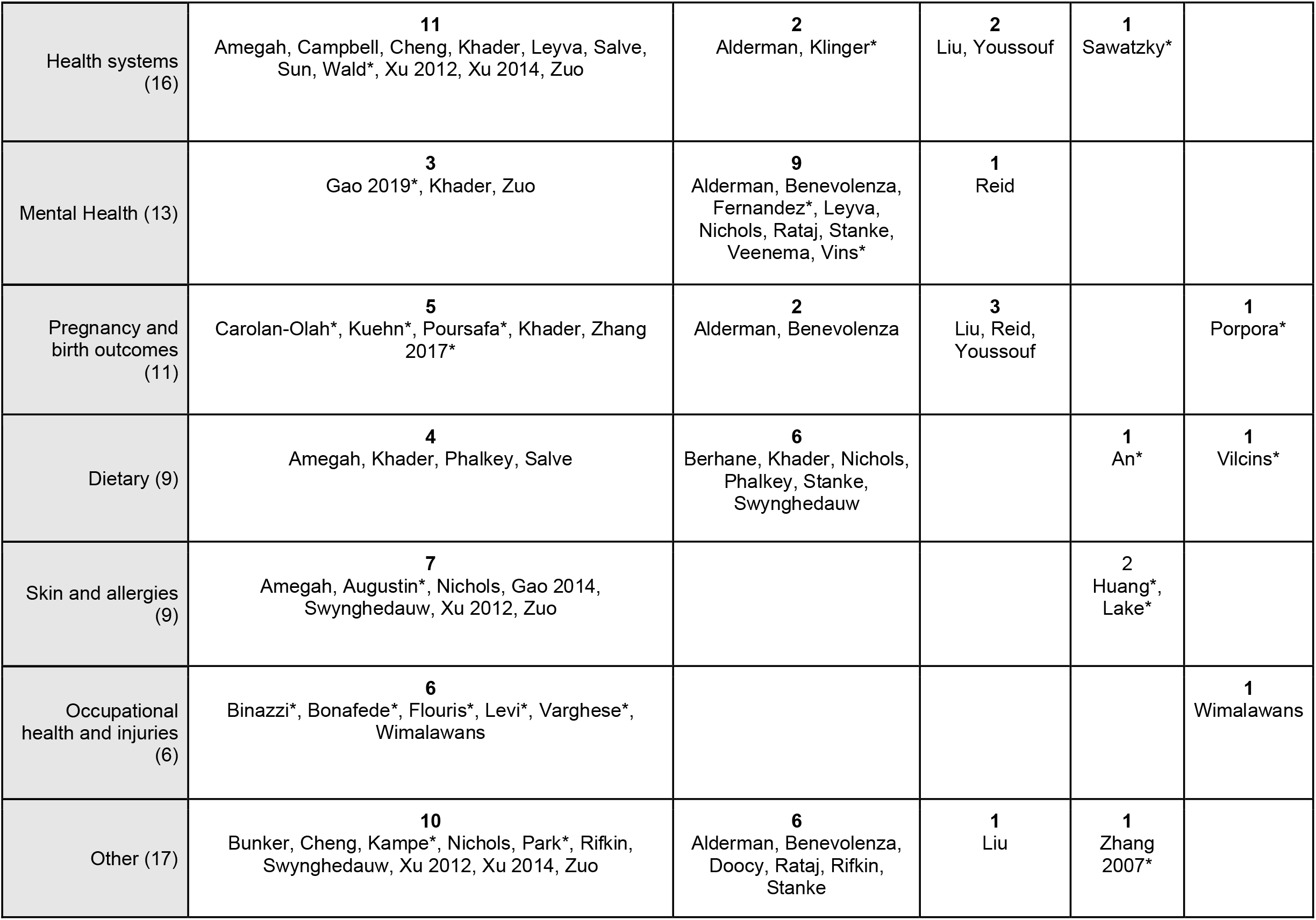

| Health Outcome | Climate Impact |  |  |  |  |
| --- | --- | --- | --- | --- | --- |
|  | Meteorological (71) | Extreme weather (24) | Air quality (7) | General (5) | Other (3) |
| Infectious diseases (41) | **35**<br>Amegah, Babaie*, Bai*, Berhane, Bernhardt*, Cheng, Coates*, de Sousa, Dhimal*, Duan*, Fan*, Gao 2014, Ghazani*, Gracia*, Hedlund, Hii*, Khader, Lal 2015*, Lal 2019*, Levy, Leyva, Li, Matysiak, Naish*, Nichols, Philipsborn*, Phung, Racloz*, Semenza*, Stensgaard, Swynghedau, Waits*, Welch*, Xu 2012, Yu 2015* | **14**<br>Alderman, Berhane, Brown*, Cann*, Hedlund, Levy, Li, Matysiak, Nichols, Phung, Stanke, Swynghedau, Tall*, Veenema |  |  |  |
| Mortality (32) | **24**<br>Amegah, Bunker, Campbell, Cheng, Cunrui*, Ghanizadeh, Hajat*, Khader, Lawton, Leyva, Lian, Moghdamnia*, Nichols, Odame*, Salve, Sanderson*, Sun, Swynghedauw, Witt, Xu 2012, Xu 2014, Xu 2016*, Yu 2012*, Zuo | **5**<br>Alderman, Doocy, Leyva, Stanke, Veenema | **5**<br>Leyva, Liu, Madniyazi*, Reid, Youssouf |  |  |
| Respiratory, cardio-vascular, cardio-pulmonary and neurological (22) | **17**<br>Amegah, Bunker, Cheng, Cong*, de Sousa, Gao 2014, Ghanizadeh, Lawton, Leyva, Lian, Nichols, Sun, Witt, Xu 2012, Xu 2014, Xu 2018*, Zuo | **1**<br>Stanke | **6**<br>Khader, Leyva, Liu, Nichols, Reid, Youssouf |  |  |
| Health systems<br>(16) | **11**<br>Amegah, Campbell, Cheng, Khader, Leyva, Salve,<br>Sun, Wald*, Xu 2012, Xu 2014, Zuo | **2**<br>Alderman, Klinger* | **2**<br>Liu, Youssouf | **1**<br>Sawatzky* |  |
| Mental Health (13) | **3**<br>Gao 2019*, Khader, Zuo | **9**<br>Alderman, Benevolenza,<br>Fernandez*, Leyva,<br>Nichols, Rataj, Stanke,<br>Veenema, Vins* | **1**<br>Reid |  |  |
| Pregnancy and<br>birth outcomes<br>(11) | **5**<br>Carolán-Olah*, Kuehn*, Poursafa*, Khader, Zhang<br>2017* | **2**<br>Alderman, Benevolenza | **3**<br>Liu, Reid,<br>Youssouf |  | **1**<br>Porpora* |
| Dietary (9) | **4**<br>Amegah, Khader, Phalkey, Salve | **6**<br>Berhane, Khader, Nichols,<br>Phalkey, Stanke,<br>Swynghedauw |  | **1**<br>An* | **1**<br>Vilcins* |
| Skin and allergies<br>(9) | **7**<br>Amegah, Augustin*, Nichols, Gao 2014,<br>Swynghedauw, Xu 2012, Zuo |  |  | **2**<br>Huang*,<br>Lake* |  |
| Occupational<br>health and injuries<br>(6) | **6**<br>Binazzi*, Bonafede*, Flouris*, Levi*, Varghese*,<br>Wimalawans |  |  |  | **1**<br>Wimalawans |
| Other (17) | **10**<br>Bunker, Cheng, Kampe*, Nichols, Park*, Rifkin,<br>Swynghedauw, Xu 2012, Xu 2014, Zuo | **6**<br>Alderman, Benevolenza,<br>Doocy, Rataj, Rifkin,<br>Stanke | **1**<br>Liu | **1**<br>Zhang<br>2007* |  |

## References

1 Portier C, Tart K, Carter S, et al. A Human Health Perspective On Climate Change A Report Outlining the Research Needs on the Human Health Effects of Climate Change. Environmental Health Perspectives and the National Institute of Environmental Health Sciences, 2010.

2 Watts N, Amann M, Arnell N, et al. The 2019 report of The Lancet Countdown on health and climate change: ensuring that the health of a child born today is not defined by a changing climate. Lancet 2019; 394: 1836–78.

3 Alderman K, Turner LR, Tong SL. Floods and human health: A systematic review. Environ Int 2012; 47: 37–47.

4 Coates SJ, Davis MDP, Andersen LK. Temperature and humidity affect the incidence of hand, foot, and mouth disease: a systematic review of the literature-a report from the International Society of Dermatology Climate Change Committee. Int J Dermatol 2019; 58: 388–99.

5 Duan C, Zhang X, Jin H, et al. Meteorological factors and its association with hand, foot and mouth disease in Southeast and East Asia areas: a meta-analysis. Epidemiology & Infection 2018; 147: 1–18.

6 Babaie J, Barati M, Azizi M, Ephtekhari A, Sadat SJ. A systematic evidence review of the effect of climate change on malaria in Iran. J Parasit Dis 2018; 42: 331–40.

7 Chua PL, Dorotan MM, Sigua JA, Estanislao RD, Hashizume M, Salazar MA. Scoping Review of Climate Change and Health Research in the Philippines: A Complementary Tool in Research Agenda-Setting. Int J Environ Res Public Health 2019; 16. DOI:10.3390/ijerph16142624.

8 Lal A, Lill AW, McIntyre M, Hales S, Baker MG, French NP. Environmental change and enteric zoonoses in New Zealand: a systematic review of the evidence. Aust N Z J Public Health 2015; 39: 63–8.

9 Li C, Lu Y, Liu J, Wu X. Climate change and dengue fever transmission in China: Evidences and challenges. Sci Total Environ 2018; 622-623: 493–501.

10 Herlihy N, Bar-Hen A, Verner G, et al. Climate change and human health: what are the research trends? A scoping review protocol. BMJ Open 2016; 6: e012022.

11 Hosking J, Campbell-Lendrum D. How well does climate change and human health research match the demands of policymakers? A scoping review. Environ Health Perspect 2012; 120: 1076–82.

12 Pollock M, Fernandes RM, Becker LA, Pieper D, Hartling L. Chapter V: Overviews of Reviews. In: Higgins Jpt, Thomas J, Chandler J, Cumpston M, Li T, Page MJ, Welch VA, ed. Cochrane Handbook for Systematic Reviews of Interventions version 6.1 (updated September 2020). Cochrane, 2020.

13 Witteman HO, Dansokho SC, Ndjaboue R, Provencher T, Poulin-Rheault RA, Poirier-Bergeron L, Beaudoin C, Fallon C, Rocque R, Tricco A. Climate change and human health: an overview of systematic reviews. 2019; published online Dec 4. https://www.crd.york.ac.uk/PROSPERO/display_record.php?RecordID=145972 (accessed Aug 8, 2020).

14 Page M, McKenzie J, Bossuyt P, Boutron I, Hoffmann TC, Mulrow C, Shamseer L, Chou R, Glanville J, Grimshaw JM, Hróbjartsson A, et al. Updating the PRISMA reporting guideline for systematic reviews and meta-analyses. 2020. DOI:10.17605/OSF.IO/P93GE.

15 Pollock M, Fernandes RM, Pieper D, et al. Preferred Reporting Items for Overviews of Reviews (PRIOR): a protocol for development of a reporting guideline for overviews of reviews of healthcare interventions. Syst Rev 2019; 8: 335.

16 About Cochrane Reviews. https://www.cochranelibrary.com/about/about-cochrane-reviews (accessed Sept 14, 2020).

17 World Health Organization. Preamble to the Constitution of WHO as adopted by the International Health Conference. New York, 19 June - 22 July 1946 signed on 22 July 1946 by the representatives of 61 States (Official Records of WHO, no. 2, p. 100) and entered into force on 7 April 1948. The definition has not been amended since 1948. https://apps.who.int/gb/bd/pdf_files/BD_49th-en.pdf#page=7.

18 Covidence systematic review software. www.covidence.org.

19 Boylan S, Beyer K, Schlosberg D, et al. A conceptual framework for climate change, health and wellbeing in NSW, Australia. Public Health Res Pract 2018; 28. DOI:10.17061/phrp2841826.

20 Leyva EWA, Beaman A, Davidson PM. Health Impact of Climate Change in Older People: An Integrative Review and Implications for Nursing. J Nurs Scholarsh 2017; 49: 670–8.

21 Khader YS, Abdelrahman M, Abdo N, et al. Climate change and health in the Eastern Mediterranean countries: a systematic review. Rev Environ Health 2015; 30: 163–81.

22 An R, Ji M, Zhang S. Global warming and obesity: a systematic review. Obes Rev 2018; 19: 150–63.

23 Phalkey RK, Aranda-Jan C, Marx S, Hofle B, Sauerborn R. Systematic review of current efforts to quantify the impacts of climate change on undernutrition. Proc Natl Acad Sci U S A 2015; 112: E4522–9.

24 Stanke C, Kerac M, Prudhomme C, Medlock J, Murray V. Health Effects of Drought: A Systematic Review of the Evidence. PLoS Curr 2013. DOI:10.1371/currents.dis.7a2cee9e980f91ad7697b570bcc4b004.

25 Vins H, Bell J, Saha S, Hess JJ. The Mental Health Outcomes of Drought: A Systematic Review and Causal Process Diagram. International Journal of Environmental Research & Public Health [Electronic Resource] 2015; 12: 13251–75.

26 Xu Z, Crooks JL, Davies JM, Khan AF, Hu W, Tong S. The association between ambient temperature and childhood asthma: a systematic review. Int J Biometeorol 2018; 62: 471–81.

27 Amegah AK, Rezza G, Jaakkola JJ. Temperature-related morbidity and mortality in Sub-Saharan Africa: A systematic review of the empirical evidence. Environ Int 2016; 91: 133– 49.

28 Odame EA, Li Y, Zheng SM, Vaidyanathan A, Silver K. Assessing Heat-Related Mortality Risks among Rural Populations: A Systematic Review and Meta-Analysis of Epidemiological Evidence. Int J Environ Res Public Health 2018; 15. DOI:10.3390/ijerph15081597.

29 Porpora MG, Piacenti I, Scaramuzzino S, Masciullo L, Rech F, Panici PB. Environmental contaminants exposure and preterm birth: A systematic review. Toxics 2019; 7. DOI:10.3390/toxics7010011.

30 Bai L, Morton LC, Liu Q. Climate change and mosquito-borne diseases in China: a review. Globalization & Health 2013; 9: 10–10.

31 Cheng J, Xu Z, Zhu R, et al. Impact of diurnal temperature range on human health: a systematic review. Int J Biometeorol 2014; 58: 2011–24.

32 Phung D, Huang C, Rutherford S, Chu C, Wang X, Nguyen M. Climate Change, Water Quality, and Water-Related Diseases in the Mekong Delta Basin: A Systematic Review. Asia Pac J Public Health 2015; 27: 265–76.

33 Klinger C, Landeg O, Murray V. Power Outages, Extreme Events and Health: A Systematic Review of the Literature from 2011-2012. PLoS Curr 2014. DOI:10.1371/currents.dis.04eb1dc5e73dd1377e05a10e9edde673.

34 Sun Z, Chen C, Xu D, Li T. Effects of ambient temperature on myocardial infarction: A systematic review and meta-analysis. Environ Pollut 2018; 241: 1106–14.

35 Berhane K, Kumie A, Samet J. Health Effects of Environmental Exposures, Occupational Hazards and Climate Change in Ethiopia: Synthesis of Situational Analysis, Needs Assessment and the Way Forward. Ethiopian Journal of Health Development 2016; 30: 50– 6.

36 Bernhardt V, Finkelmeier F, Verhoff MA, Amendt J. Myiasis in humans-a global case report evaluation and literature analysis. Parasitol Res 2019; 118: 389–97.

37 de Sousa Tcm, Amancio F, Hacon SS, Barcellos C. [Climate-sensitive diseases in Brazil and the world: systematic review] Enfermedades sensibles al clima en Brasil y el mundo: revision sistematica. Rev Panam Salud Publica 2018; 42: e85.

38 Dhimal M, Ahrens B, Kuch U. Climate Change and Spatiotemporal Distributions of Vector-Borne Diseases in Nepal--A Systematic Synthesis of Literature. PLoS ONE [Electronic Resource] 2015; 10: e0129869.

39 Fan J, Wei W, Bai Z, et al. A systematic review and meta-analysis of dengue risk with temperature change. Int J Environ Res Public Health 2015; 12: 1–15.

40 Gracia JR, Schumann B, Seidler A. Climate Variability and the Occurrence of Human Puumala Hantavirus Infections in Europe: A Systematic Review. Zoonoses Public Health 2015; 62: 465–78.

41 Hedlund C, Blomstedt Y, Schumann B. Association of climatic factors with infectious diseases in the Arctic and subarctic region--a systematic review. Glob Health Action 2014; 7: 24161.

42 Hii YL, Zaki RA, Aghamohammadi N, Rocklov J. Research on Climate and Dengue in Malaysia: A Systematic Review. Current Environmental Health Reports 2016; 3: 81–90.

43 Matysiak A, Roess A. Interrelationship between Climatic, Ecologic, Social, and Cultural Determinants Affecting Dengue Emergence and Transmission in Puerto Rico and Their Implications for Zika Response. J Trop Med 2017; 2017. DOI:10.1155/2017/8947067.

44 Naish S, Dale P, Mackenzie JS, McBride J, Mengersen K, Tong S. Climate change and dengue: a critical and systematic review of quantitative modelling approaches. BMC Infect Dis 2014; 14: 167–167.

45 Nichols A, Maynard V, Goodman B, Richardson J. Health, climate change and sustainability: a systematic review and thematic analysis of the literature. Environ Health Insights 2009; : 63–88.

46 Racloz V, Ramsey R, Tong S, Hu W. Surveillance of dengue fever virus: a review of epidemiological models and early warning systems. PLoS Neglected Tropical Diseases [electronic resource] 2012; 6: e1648.

47 Swynghedauw B. [Medical consequences of global warming]. Presse Med 2009; 38: 551–61.

48 Waits A, Emelyanova A, Oksanen A, Abass K, Rautio A. sHuman infectious diseases and the changing climate in the Arctic. Environ Int 2018; 121: 703–13.

49 Xu Z, Etzel RA, Su H, Huang C, Guo Y, Tong S. Impact of ambient temperature on children’s health: a systematic review. Environ Res 2012; 117: 120–31.

50 Yu W, Mengersen K, Dale P, et al. Projecting Future Transmission of Malaria Under Climate Change Scenarios: Challenges and Research Needs. Crit Rev Environ Sci Technol 2015; 45: 777–811.

51 Veenema TG, Thornton CP, Lavin RP, Bender AK, Seal S, Corley A. Climate Change-Related Water Disasters’ Impact on Population Health. J Nurs Scholarsh 2017; 49: 625–34.

52 Brown L, Murray V. Examining the relationship between infectious diseases and flooding in Europe: A systematic literature review and summary of possible public health interventions. Disaster Health 2013; 1: 117–27.

53 Tall JA, Gatton ML, Tong S. Ross River Virus Disease Activity Associated With Naturally Occurring Nontidal Flood Events in Australia: A Systematic Review. J Med Entomol 2014; 51: 1097–108.

54 Ghazani M, FitzGerald G, Hu WB, Toloo G, Xu ZW. Temperature Variability and Gastrointestinal Infections: A Review of Impacts and Future Perspectives. Int J Environ Res Public Health 2018; 15. DOI:10.3390/ijerph15040766.

55 Gao J, Sun Y, Lu Y, Li L. Impact of ambient humidity on child health: a systematic review. PLoS ONE [Electronic Resource] 2014; 9: e112508.

56 Lal A, Fearnley E, Wilford E. Local weather, flooding history and childhood diarrhoea caused by the parasite Cryptosporidium spp.: A systematic review and meta-analysis. Sci Total Environ 2019; 674: 300–6.

57 Levy K, Woster AP, Goldstein RS, Carlton EJ. Untangling the Impacts of Climate Change on Waterborne Diseases: a Systematic Review of Relationships between Diarrheal Diseases and Temperature, Rainfall, Flooding, and Drought. Environ Sci Technol 2016; 50: 4905–22.

58 Philipsborn R, Ahmed SM, Brosi BJ, Levy K. Climatic Drivers of Diarrheagenic Escherichia coli Incidence: A Systematic Review and Meta-analysis. J Infect Dis 2016; 214: 6–15.

59 Semenza JC, Herbst S, Rechenburg A, et al. Climate change impact assessment of food-and waterborne diseases. Crit Rev Environ Sci Technol 2012; 42: 857–90.

60 Stensgaard AS, Vounatsou P, Sengupta ME, Utzinger J. Schistosomes, snails and climate change: Current trends and future expectations. Acta Trop 2019; 190: 257–68.

61 Welch K, Shipp-Hilts A, Eidson M, Saha S, Zansky S. Salmonella and the changing environment: systematic review using New York State as a model. J Water Health 2019; 17: 179–95.

62 Cann KF, Thomas DR, Salmon RL, Wyn-Jones AP, Kay D. Extreme water-related weather events and waterborne disease. Epidemiology & Infection 2013; 141: 671–86.

63 Bunker A, Wildenhain J, Vandenbergh A, et al. Effects of Air Temperature on Climate-Sensitive Mortality and Morbidity Outcomes in the Elderly; a Systematic Review and Meta-analysis of Epidemiological Evidence. EBioMedicine 2016; 6: 258–68.

64 Campbell S, Remenyi TA, White CJ, Johnston FH. Heatwave and health impact research: A global review. Health Place 2018; 53: 210–8.

65 Cunrui H, Barnett AG, Xiaoming W, Vaneckova P, FitzGerald G, Shilu T. Projecting Future Heat-Related Mortality under Climate Change Scenarios: A Systematic Review. Environ Health Perspect 2011; 119: 1681–90.

66 Ghanizadeh G, Heidari M, Seifi B, Jafari H, Pakjouei S. The effect of climate change on cardiopulmonary disease-a systematic review. J Clin Diagn Res 2017; 11: IE01–4.

67 Hajat S, Kosatky T. Heat-related mortality: a review and exploration of heterogeneity. Journal of Epidemiology & Community Health 2010; 64: 753–60.

68 Lawton EM, Pearce H, Gabb GM. Review article: Environmental heatstroke and long-term clinical neurological outcomes: A literature review of case reports and case series 2000– 2016. Emerg Med Australas 2019; 31: 163–73.

69 Lian H, Ruan YP, Liang RJ, Liu XL, Fan ZJ. Short-Term Effect of Ambient Temperature and the Risk of Stroke: A Systematic Review and Meta-Analysis. Int J Environ Res Public Health 2015; 12: 9068–88.

70 Moghadamnia MT, Ardalan A, Mesdaghinia A, Keshtkar A, Naddafi K, Yekaninejad MS. Ambient temperature and cardiovascular mortality: A systematic review and meta-analysis. PeerJ 2017; 2017: 3574.

71 Salve HR, Parthasarathy R, Krishnan A, Pattanaik DR. Impact of ambient air temperature on human health in India. Rev Environ Health 2018; 33: 433–9.

72 Sanderson M, Arbuthnott K, Kovats S, Hajat S, Falloon P. The use of climate information to estimate future mortality from high ambient temperature: A systematic literature review. PLoS ONE [Electronic Resource] 2017; 12: e0180369.

73 Witt C, Schubert AJ, Jehn M, et al. The Effects of Climate Change on Patients With Chronic Lung Disease. A Systematic Literature Review. Dtsch Arztebl Int 2015; 112: 878– 83.

74 Xu Z, Sheffield PE, Su H, Wang X, Bi Y, Tong S. The impact of heat waves on children’s health: a systematic review. Int J Biometeorol 2014; 58: 239–47.

75 Xu Z, FitzGerald G, Guo Y, Jalaludin B, Tong S. Impact of heatwave on mortality under different heatwave definitions: A systematic review and meta-analysis. Environ Int 2016; 89-90: 193–203.

76 Yu W, Mengersen K, Wang X, et al. Daily average temperature and mortality among the elderly: a meta-analysis and systematic review of epidemiological evidence. Int J Biometeorol 2012; 56: 569–81.

77 Doocy S, Dick A, Daniels A, Kirsch TD. The Human Impact of Tropical Cyclones: A Historical Review of Events 1980-2009 and Systematic Literature Review. PLoS Curr 2013. DOI:10.1371/currents.dis.2664354a5571512063ed29d25ffbce74.

78 Madaniyazi L, Guo Y, Yu W, Tong S. Projecting future air pollution-related mortality under a changing climate: Progress, uncertainties and research needs. Environ Int 2015; 75: 21–32.

79 Liu JC, Pereira G, Uhl SA, Bravo MA, Bell ML. A systematic review of the physical health impacts from non-occupational exposure to wildfire smoke. Environ Res 2015; 136: 120– 32.

80 Reid CE, Brauer M, Johnston FH, Jerrett M, Balmes JR, Elliott CT. Critical Review of Health Impacts of Wildfire Smoke Exposure. Environ Health Perspect 2016; 124: 1334–43.

81 Youssouf H, Liousse C, Roblou L, et al. Non-accidental health impacts of wildfire smoke. International Journal of Environmental Research & Public Health [Electronic Resource] 2014; 11: 11772–804.

82 Cong XW, Xu XJ, Zhang YL, Wang QH, Xu L, Huo X. Temperature drop and the risk of asthma: a systematic review and meta-analysis. Environ Sci Pollut Res 2017; 24: 22535–46.

83 Zuo J, Pullen S, Palmer J, Bennetts H, Chileshe N, Ma T. Impacts of heat waves and corresponding measures: a review. J Clean Prod 2015; 92: 1–12.

84 Sawatzky A, Cunsolo A, Jones-Bitton A, Middleton J, Harper SL. Responding to Climate and Environmental Change Impacts on Human Health via Integrated Surveillance in the Circumpolar North: A Systematic Realist Review. International Journal of Environmental Research & Public Health [Electronic Resource] 2018; 15: 30.

85 Wald A. Emergency Department Visits and Costs for Heat-Related Illness Due to Extreme Heat or Heat Waves in the United States: An Integrated Review. Nurs Econ 2019; 37: 35– 48.

86 Gao JJ, Cheng Q, Duan J, et al. Ambient temperature, sunlight duration, and suicide: A systematic review and meta-analysis. Sci Total Environ 2019; 646: 1021–9.

87 Benevolenza MA, DeRigne L. The impact of climate change and natural disasters on vulnerable populations: A systematic review of literature. J Hum Behav Soc Environ 2019; 29: 266–81.

88 Rataj E, Kunzweiler K, Garthus-Niegel S. Extreme weather events in developing countries and related injuries and mental health disorders - a systematic review. BMC Public Health 2016; 16: 1020–1020.

89 Fernandez A, Black J, Jones M, et al. Flooding and Mental Health: A Systematic Mapping Review. PLoS ONE [Electronic Resource] 2015; 10. DOI:10.1371/journal.pone.0119929.

90 Carolan-Olah M, Frankowska D. High environmental temperature and preterm birth: A review of the evidence. Midwifery 2014; 30: 50–9.

91 Kuehn L, McCormick S. Heat Exposure and Maternal Health in the Face of Climate Change. International Journal of Environmental Research & Public Health [Electronic Resource] 2017; 14: 29.

92 Poursafa P, Keikha M, Kelishadi R. Systematic review on adverse birth outcomes of climate change. J Res Med Sci 2015; 20: 397–402.

93 Zhang YQ, Yu CH, Wang L. Temperature exposure during pregnancy and birth outcomes: An updated systematic review of epidemiological evidence. Environ Pollut 2017; 225: 700– 12.

94 Vilcins D, Sly PD, Jagals P. Environmental Risk Factors Associated with Child Stunting: A Systematic Review of the Literature. Annals of Global Health 2018; 84: 551–62.

95 Huang KC, Weng HH, Yang TY, Chang TS, Huang TW, Lee MS. Distribution of Fatal Vibrio Vulnificus Necrotizing Skin and Soft-Tissue Infections: A Systematic Review and Meta-Analysis. Medicine 2016; 95: e2627.

96 Lake IR, Jones NR, Agnew M, et al. Climate Change and Future Pollen Allergy in Europe. Environ Health Perspect 2017; 125: 385–91.

97 Augustin J, Franzke N, Augustin M, Kappas M. Does climate change affect the incidence of skin and allergic diseases in Germany? J Dtsch Dermatol Ges 2008; 6: 632–8.

98 Binazzi A, Levi M, Bonafede M, et al. Evaluation of the impact of heat stress on the occurrence of occupational injuries: Meta-analysis of observational studies. Am J Ind Med 2019; 62: 233–43.

99 Bonafede M, Marinaccio A, Asta F, Schifano P, Michelozzi P, Vecchi S. The association between extreme weather conditions and work-related injuries and diseases. A systematic review of epidemiological studies. Annali Dell Istituto Superiore Di Sanita 2016; 52: 357– 67.

100 Flouris AD, Dinas PC, Ioannou LG, et al. Workers’ health and productivity under occupational heat strain: a systematic review and meta-analysis. The lancet Planetary Health 2018; 2: e521–31.

101 Levi M, Kjellstrom T, Baldasseroni A. Impact of climate change on occupational health and productivity: a systematic literature review focusing on workplace heat. Medicina del Lavoro 2018; 109: 163–79.

102 Varghese BM, Hansen A, Bi P, Pisaniello D. Are workers at risk of occupational injuries due to heat exposure? A comprehensive literature review. Saf Sci 2018; 110: 380–92.

103 Wimalawansa SA, Wimalawansa SJ. Environmentally induced, occupational diseases with emphasis on chronic kidney disease of multifactorial origin affecting tropical countries. Annals of Occupational and Environmental Medicine 2016; 28. DOI:10.1186/s40557-016-0119-y.

104 Zhang Y, Bi P, Hiller JE. Climate change and disability -- adjusted life years. J Environ Health 2007; 70: 32–6.

105 Park KY, Kim HJ, Ahn HS, Yim SY, Jun JB. Association between acute gouty arthritis and meteorological factors: An ecological study using a systematic review and meta-analysis. Semin Arthritis Rheum 2017; 47: 369–75.

106 Kampe EOI, Kovats S, Hajat S. Impact of high ambient temperature on unintentional injuries in high-income countries: a narrative systematic literature review. BMJ Open 2016; 6. DOI:10.1136/bmjopen-2015-010399.

107 Rifkin DI, Long MW, Perry MJ. Climate change and sleep: A systematic review of the literature and conceptual framework. Sleep Med Rev 2018; 42: 3–9.

108 Clayton S. Climate anxiety: Psychological responses to climate change. J Anxiety Disord 2020; 74: 102263.

109 Davenport L. Emotional Resiliency in the Era of Climate Change: A Clinician’s Guide. London: Jessica Kingsley Publishers, 2017.

110 Maibach EW, Nisbet M, Baldwin P, Akerlof K, Diao G. Reframing climate change as a public health issue: an exploratory study of public reactions. BMC Public Health 2010; 10: 299.

111 Stoknes PE. What we think about when we try not to think about global warming: Toward a new psychology of climate action. White River Junction, Vermont: Chelsea Green Publishing 2015.

112 Costello A, Montgomery H, Watts N. Climate change: the challenge for healthcare professionals. BMJ 2013; 347: f6060.

113 Yang L, Liu C, Hess J, Phung D, Huang C. Health professionals in a changing climate: protocol for a scoping review. BMJ Open 2019; 9: e024451.

